# Projections of wastewater as an indicator of COVID-19 cases in corrections facilities: a modelling study

**DOI:** 10.1101/2023.10.31.23296864

**Authors:** Dan Han, Pamela Linares, Rochelle H. Holm, Kartik Chandran, Ted Smith

## Abstract

**Background:** Although prison facilities are not fully isolated from the communities they are located within, the majority of the population is confined and requires high levels of health vigilance and protection. This study sought to examine the dynamic relationship between facility level wastewater viral RNA concentration and probability of at least one positive COVID-19 case within the facility.

**Methods:** The study period was January 11, 2021 through May 12, 2023. Wastewater samples were collected and analyzed for SARS-CoV-2 (N1) and pepper mild mottle virus (PMMoV) three times per week across 14 prison facilities in Kentucky (USA). Confirmed positive clinical case reports were also provided. A hierarchical Bayesian spatial-temporal model with a latent lagged process was developed.

**Findings:** We modeled a facility-specific SARS-CoV-2 (N1) normalized by PMMoV wastewater ratio associated with at least one COVID-19 facility case with an 80% probability. The ratio differs among facilities. Across the 14 facilities, our model demonstrates an average capture rate of 94·95% via the N1/PMMoV threshold with *p*_*ts*_ ≥ 0·5. However, it is noteworthy as the *p*_*ts*_ threshold is set higher, such as at 0·9 or above, the model’s average capture rate reduces to 60%. This robust performance underscores the model’s effectiveness in accurately detecting the presence of positive COVID-19 cases of incarcerated people.

**Interpretation:** The findings of this study provide a correction facility-specific threshold model for public health response based on frequent wastewater surveillance.

## 1. Introduction

Wastewater-based epidemiology (WBE) at a community level typically involves an associated amount of SARS-CoV-2 present in wastewater with confirmed COVID-19 cases.^1–6^ Yet, the amount of SARS-CoV-2 virus shed into wastewater can vary among infected individuals, depending on factors such as disease severity and individual differences.^7^ Correctional and detention facilities in the United States have reported over 900,000 cases of COVID-19 among incarcerated persons and staff, and with nearly 3,500 deaths.^8,9^ Although prison facilities are not fully isolated from the communities they are located within, the majority of the population is not transient and requires high levels of health vigilance and protection.^10^ There have been few examples of single or a few clusters of corrections facility testing alongside large community scale SARS-CoV-2 monitoring.^11–13^ Corrections facilities are also unique from a health equity angle, as healthcare access and testing are universally available.

We present the development of a hierarchical Bayesian spatial-temporal model within corrections facilities. In contrast to extant count time series models,^14–18^ the present model not only accounts for potential instances of non-reports but also explicitly incorporates the dynamic temporal lags between wastewater-based virus concentration and the occurrence of positive clinical cases. This model aims to provide insights and answers to the wastewater concentration associated with the detection of at least one positive COVID-19 case of an incarcerated person. The findings may contribute to the development of guidelines and proactive wastewater guided decision-making and planning for public health authorities at corrections facilities, policymakers, and healthcare systems.

## 2. Methods

### 2.1 Wastewater sample collection and analysis

Wastewater samples were collected three times per week using a 24-h composite sample at 14 facilities from January 11, 2021 through May 12, 2023. The sample collection location was a utility access hole outside the main facility and dedicated to the influent from the facility, offering ease of access for the sampling personnel external to physical security barriers. Samples were transported on ice to Eurofins Microbiology Laboratories, Inc. (Louisville, KY, USA) for analysis. Samples were processed within 24 hours of collection, concentrated overnight, and quantified in triplicate by reverse transcription polymerase chain reaction (RT-PCR). Data for SARS-CoV-2 (N1) and pepper mild mottle virus (PMMoV) targets were reported on an unconcentrated sample basis (copies/ml of wastewater). The detection limit for SARS-CoV-2 (N1) was 20 copies/mL and for PMMoV was 100 copies/mL. We used the weekly average wastewater ratios.

### 2.2 Clinical testing data

De-identified clinical testing positive case counts were provided by Kentucky Department of Corrections for the sampling period for both staff and inmates. Rapid tests were done first at incarcerated person intake, and positive results were further confirmed via polymerase chain reaction (PCR) testing for COVID-19. Most facilities conducted census testing of typically a random 10% of the population weekly in additional to the testing of those who presented in clinic with symptoms. Occupational testing of staff occurred under a range of processes that varied by facility and over time. We only consider PCR clinical data in this analysis. These data contained information concerning: the number of PCR positive clinical cases and total number of tests per facility separated by incarcerated persons and staff; the report date; and facility name.

### 2.3 Facility population and capacity data

De-identified number of daily people incarcerated and facility operational capacity data were provided by Kentucky Department of Corrections for the sampling period. These data contained information concerning: the number of incarcerated people per facility; the report date; and the facility name. We used the weekly average inmate population in the 14 facilities ranging from 170 to 1604 (Supplemental Material Table A.1).

### 2.4 Statistical Analysis

Full model details are provided in Supplemental Material Appendix B. In brief, our model is applied to the counts of positive inmate cases in 14 correctional facilities paired with wastewater data covering the period from January 11, 2021 to May 12, 2023. We assume for each facility *s,s* = 1,2, …,*J*, has independent reporting probabilities π_*ts*_ at time *t*. π_*ts*_ is determined by dividing the total current population by the facility’s operational capacity (population ratio) and the positive employee number 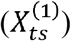. We denote the observed number of positive cases in facility *s* at time *t* as *Z*_*ts*_. For the true number of positive cases (*N*_*ts*_) in facility s at time *t*, we employ a latent Poisson process with parameter *λ*_*ts*_ which follows Gamma distribution with shape parameter *h* and scale parameter 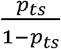. The parameter *p*_*ts*_ represents the probability of at least one positive case (*P*(*N*_*ts*_ ≥ 1)) and is influenced by the SARS-CoV-2 (N1)/PMMoV ratio. Previous studies have reported wide variability in wastewater lead times when compared to case data.^19^ We use an autoregressive latent process *γ*_*ts*_ to capture the lag effects in facility *s* at time *t*. In summary, the Bayesian hierarchical spatial temporal model has the following structure:

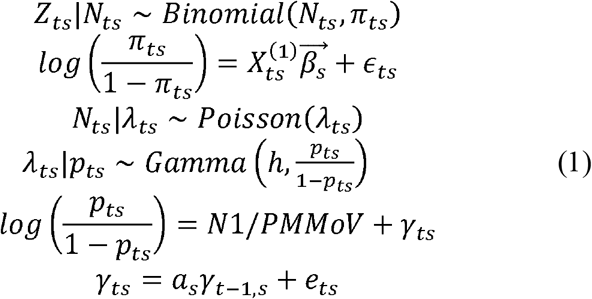

with notations:

*Z*_*ts*_ The recorded/observed positive case number in facility *s* at time *t*

*N*_*ts*_ The true positive case number in facility *s* at time *t*

*π*_*ts*_ Reporting probability in facility *s* at time *t*

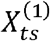 The matrix of variables influencing reporting probability

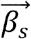 The coefficient vector for variables influencing reporting probability in facility *s*

*ϵ*_*ts*_ Normal distributed error term in facility *s* at time *t*

*λ*_*ts*_ Intensity parameter for Poisson process *N*_*ts*_ in facility *s* at time *t*

*p*_*ts*_ The probability of least one positive case (*P*(*N*_*ts*_ ≥ 1)) in facility *s* at time *t*

*h* Shape parameter of Gamma distribution

*γ*_*ts*_ The latent process that captures the lag effects in facility *s* at time *t*

*γ*_*t-1, s*_ The latent process that captures the lag effects in facility *s* at time *t*-1

*a*_*s*_ The parameter in the autoregressive model for *γ*_*ts*_ in facility *s*

*e*_*ts*_ Normal distributed error term in facility *s* at time *t*

We derived a Gibbs sampling algorithm for this Bayesian hierarchical model, enabling the data sources to inform the model within a single robust statistical framework.

This approach enabled us to calculate point estimates and Bayesian 95% confidence intervals for the spatial-temporal probability of at least one positive case (*p*_*ts*_) in the facility. This criterion value of one case was determined to be useful for communicating risk from wastewater samples for ongoing monitoring. Importantly, we identified the SARS-CoV-2 (N1)/PMMoV ratio associated with a probability of at least one positive case exceeding *p*_*ts*_ of 0·8. Furthermore, taking into account controlled *p*_*ts*_, we established a temporal-spatial alert signal when the observed SARS-CoV-2 (N1)/PMMoV ratio surpasses the predefined threshold.

The analyses were performed using R (version 4.3.0).

### 2.5 Ethics

The University of Louisville Institutional Review Board classified this project as Non-Human Subject Research (reference #: 714006).

## 3. Results and Discussion

### 3.1 Temporal and Spatial Variability of SARS-CoV-2 (N1)/PMMoV ratio

Based on the wastewater SARS-CoV-2 (N1)/PMMoV ratio, we computed the probability (*p*_*ts*_ = *P*(*N*_*ts*_ ≥ 1)) of having at least one positive case in each facility at different time points (*t* = 1,2,3,…*N*) and facility (*s* = 1,2,3,…14). In locations where a large number of cases are reported weekly, the corresponding *p*_*ts*_ values tend to be higher, indicating an increased probability of detecting at least one positive case (Figure 1). On a facility specific level, for the week where we have maximum reported cases, the estimated *p*_*ts*_ are each greater than 0·9 (Table 1; Supplemental Material Figure A.2).

**Table 1:** The weekly maximum reported positive COVID-19 cases of an incarcerated person per facility and the week in which it was observed, the associated wastewater SARS-CoV-2 (N1) copies per ml normalized by pepper mild mottle virus (PMMoV) copies per ml as a ratio, and the estimated probability (*p*_*ts*_) associated with at least one positive case.

| Facility | Week<br>(year/month/date)<br>(week number) | Maximum<br>weekly<br>clinical<br>cases<br>($Z_{ts}$) | Associated<br>wastewater<br>SARS-CoV-2<br>(N1)/PMMoV<br>ratio | Estimated<br>$p_{ts}$ |
| --- | --- | --- | --- | --- |
| A | 2023/02/19 - 2023/02/25 (108) | 29 | $1.33 \times 10^{-3}$ | 0.9703 |
| B | 2022/01/23 - 2023/01/29 (50) | 21 | $1.23 \times 10^{-3}$ | 0.9459 |
| C | 2022/01/09 - 2022/01/15 (50) | 339 | $1.31 \times 10^{-2}$ | 1 |
| D | 2022/01/23 - 2022/01/29 (52) | 166 | $8.36 \times 10^{-3}$ | 0.9993 |
| E | 2022/01/16 - 2022/01/22 (51) | 49 | $1.58 \times 10^{-3}$ | 0.9746 |
| F | 2021/03/07 - 2021/03/13 (6) | 226 | $8.94 \times 10^{-3}$ | 0.9996 |
| G | 2022/02/13 - 2022/02/19 (55) | 75 | $3.50 \times 10^{-4}$ | 0.972 |
| H | 2022/02/13 - 2022/02/19 (55) | 153 | $1.12 \times 10^{-3}$ | 0.9875 |
| I | 2022/01/30 - 2022/02/05 (53) | 135 | $1.31 \times 10^{-2}$ | 1 |
| J | 2022/01/23 - 2022/01/29 (52) | 131 | $1.60 \times 10^{-3}$ | 0.9861 |
| K | 2022/01/23 - 2022/01/29 (52) | 421 | $2.35 \times 10^{-3}$ | 0.9957 |
| L | 2022/01/30 - 2022/02/05 (53) | 180 | $2.92 \times 10^{-3}$ | 0.9903 |
| M | 2022/07/24 - 2022/07/30 (78) | 131 | $4.33 \times 10^{-3}$ | 0.9928 |
| N | 2021/02/21 - 2021/02/27 (4) | 200 | $1.20 \times 10^{-3}$ | 0.9875 |

**Figure 1:**
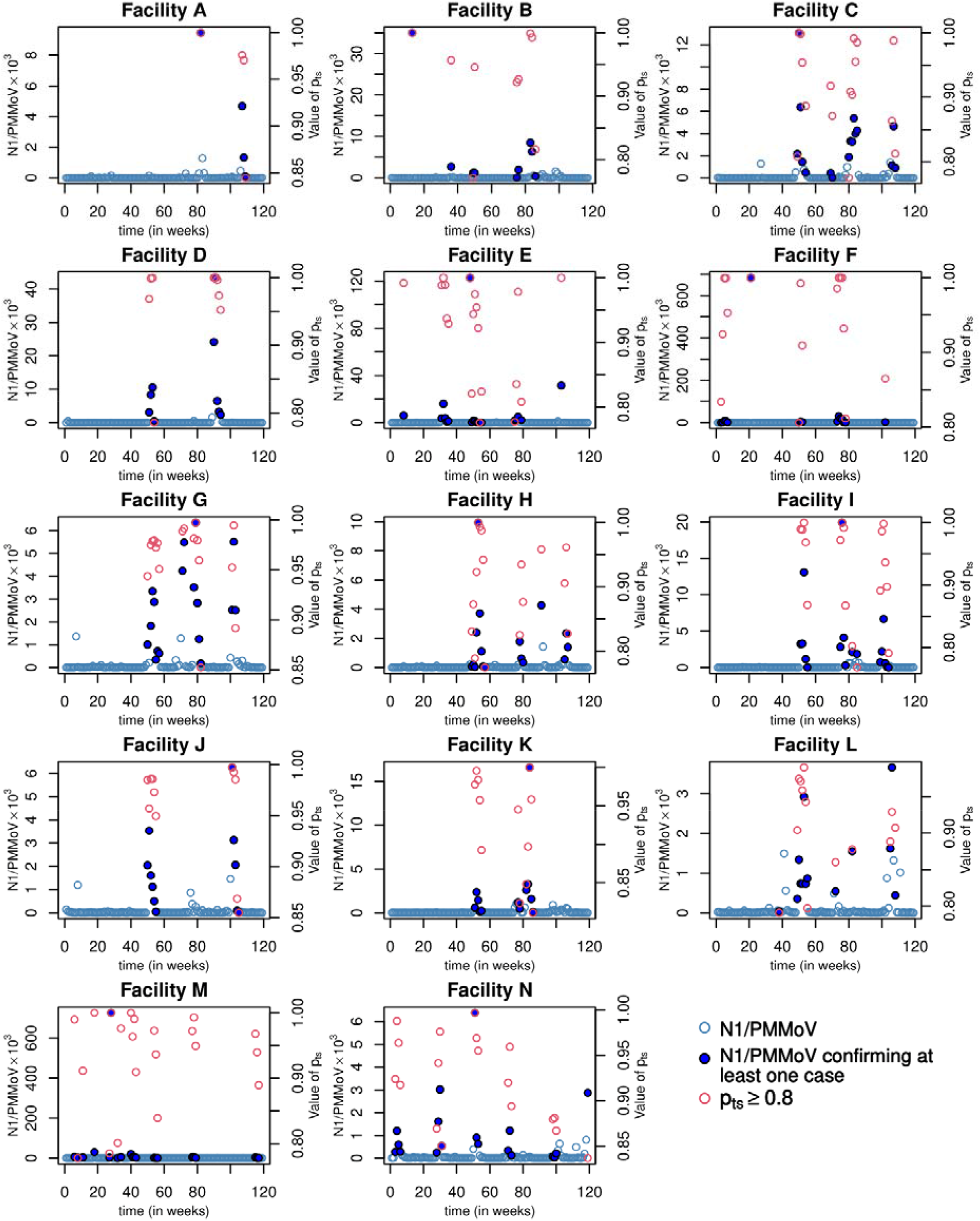
Temporal and spatial variability of SARS-CoV-2 (N1) copies per ml normalized by pepper mild mottle virus (PMMoV) copies per ml as a ratio compared to 80% probability of at least one positive COVID-19 case of an incarcerated person. Light blue outline circle indicates the wastewater SARS-CoV-2 (N1) copies per ml normalized by pepper mild mottle virus (PMMoV) copies per ml concentration ratio with no associated clinical case. A solid dark blue circle indicates the wastewater ratio associated with at least one clinical case with *P*_*ts*_ ≥ 0 · 8. The corresponding *p*_*ts*_ are plotted in red outline circle.

### 3.2 Alert Signal

The SARS-CoV-2 (N1)/PMMoV ratio corresponding to at least one case with 0·8 probability differs among facilities (Table 2).

**Table 2:** Threshold ratio per facility indicative of a high likelihood of positive cases for incarcerated people. Threshold is based on the estimated wastewater SARS-CoV-2 (N1) copies per ml normalized by pepper mild mottle virus (PMMoV) copies per ml as a ratio by facility at 80% probability of at least one positive COVID-19 case.

| Facility | SARS-CoV-2 (N1)/PMMoV ratio | | | Mean of<br>$p_{ts} \geq 0.8$ |
| --- | --- | --- | --- | --- |
|  | Minimum | Median | Mean |  |
| A | $9.11 \times 10^{-5}$ | $3.00 \times 10^{-3}$ | $3.89 \times 10^{-3}$ | 0.9476 |
| B | $6.06 \times 10^{-5}$ | $1.97 \times 10^{-3}$ | $6.38 \times 10^{-3}$ | 0.9256 |
| C | $9.55 \times 10^{-6}$ | $2.70 \times 10^{-3}$ | $3.29 \times 10^{-3}$ | 0.9137 |
| D | $5.21 \times 10^{-4}$ | $6.51 \times 10^{-3}$ | $1.14 \times 10^{-2}$ | 0.964 |
| E | $3.15 \times 10^{-5}$ | $1.94 \times 10^{-3}$ | $1.07 \times 10^{-2}$ | 0.9184 |
| F | $8.24 \times 10^{-6}$ | $4.28 \times 10^{-3}$ | $4.95 \times 10^{-2}$ | 0.9383 |
| G | $1.80 \times 10^{-4}$ | $2.53 \times 10^{-3}$ | $2.66 \times 10^{-3}$ | 0.9627 |
| H | $3.42 \times 10^{-5}$ | $8.77 \times 10^{-4}$ | $1.81 \times 10^{-3}$ | 0.8999 |
| I | $6.69 \times 10^{-6}$ | $2.16 \times 10^{-3}$ | $3.67 \times 10^{-3}$ | 0.9244 |
| J | $1.80 \times 10^{-5}$ | $1.60 \times 10^{-3}$ | $1.86 \times 10^{-3}$ | 0.9578 |
| K | $8.48 \times 10^{-5}$ | $1.30 \times 10^{-3}$ | $2.53 \times 10^{-3}$ | 0.9242 |
| L | $3.82 \times 10^{-5}$ | $7.33 \times 10^{-4}$ | $1.19 \times 10^{-3}$ | 0.9072 |
| M | $9.03 \times 10^{-5}$ | $3.41 \times 10^{-3}$ | $3.91 \times 10^{-2}$ | 0.9316 |
| N | $2.68 \times 10^{-5}$ | $5.69 \times 10^{-4}$ | $1.14 \times 10^{-3}$ | 0.9213 |

Assuming a controlled probability of at least one confirmed case existing at time t in location s, with a minimum threshold 0·8, *p*_*ts*_ ≥ 0·8, we can determine the minimum temporal dynamical SARS-CoV-2 N1/PMMoV ratio which might be informative for health leadership at the facility:

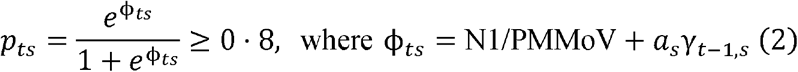

we get:

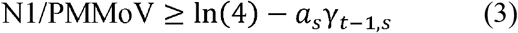

If ln(4) - *a*_*s*_γ_*t*−1,*s*_ < 0, this implies regardless of the SARS-CoV-2 (N1)/PMMoV ratio at time t in location s, the ratio for the previous week (at time *t* − 1) is sufficiently high to indicate the presence of a positive case at the current time *t*. This observation highlights the significance of considering the previous week’s SARS-CoV-2 (N1)/PMMoV wastewater ratio in determining the likelihood of positive clinical case detection.^19^

By incorporating this threshold ratio, we can effectively identify situations where the previous week’s SARS-CoV-2 (N1)/PMMoV ratio sufficiently indicates the presence of positive cases at the current time. By monitoring and analyzing the temporal dynamics of the SARS-CoV-2 (N1)/PMMoV ratio alert signal in wastewater samples, if the observed value exceeds the determined threshold, an alert can be raised, signaling the likelihood of at least one confirmed case within the system.

### 3.3 Sensitivity

However, it is noteworthy as the *p*_*ts*_ threshold is set even higher, such as at 0·9 or above, the model’s average capture rate reduces to 60%. This decline highlights a potential trade-off between leveraging higher positive case existence probabilities to filter SARS-CoV-2 (N1)/PMMoV ratio levels and the risk of missing instances where only a single positive case is reported during a given week, often constrained by the laboratory’s detection limit or turn around time. The appropriate threshold of *p*_*ts*_ is context-dependent and varies based on the specific spatial location. For instance, as shown in Table 3, when *p*_*ts*_ =0 ·8, facilities A, C, E, F, G, H, I and J have a capture ratio exceeding 80%, while facilities B, D, K, M, and N maintain a capture ratio of more than 60% but below 80%. Location L exhibits the lowest performance with a capture ratio of only 41·3%. However, adjusting the threshold to *p*_*ts*_ =0·5 leads to a significant enhancement in capture ratios; for example, location L improves to 95·2%.

**Table 3:**
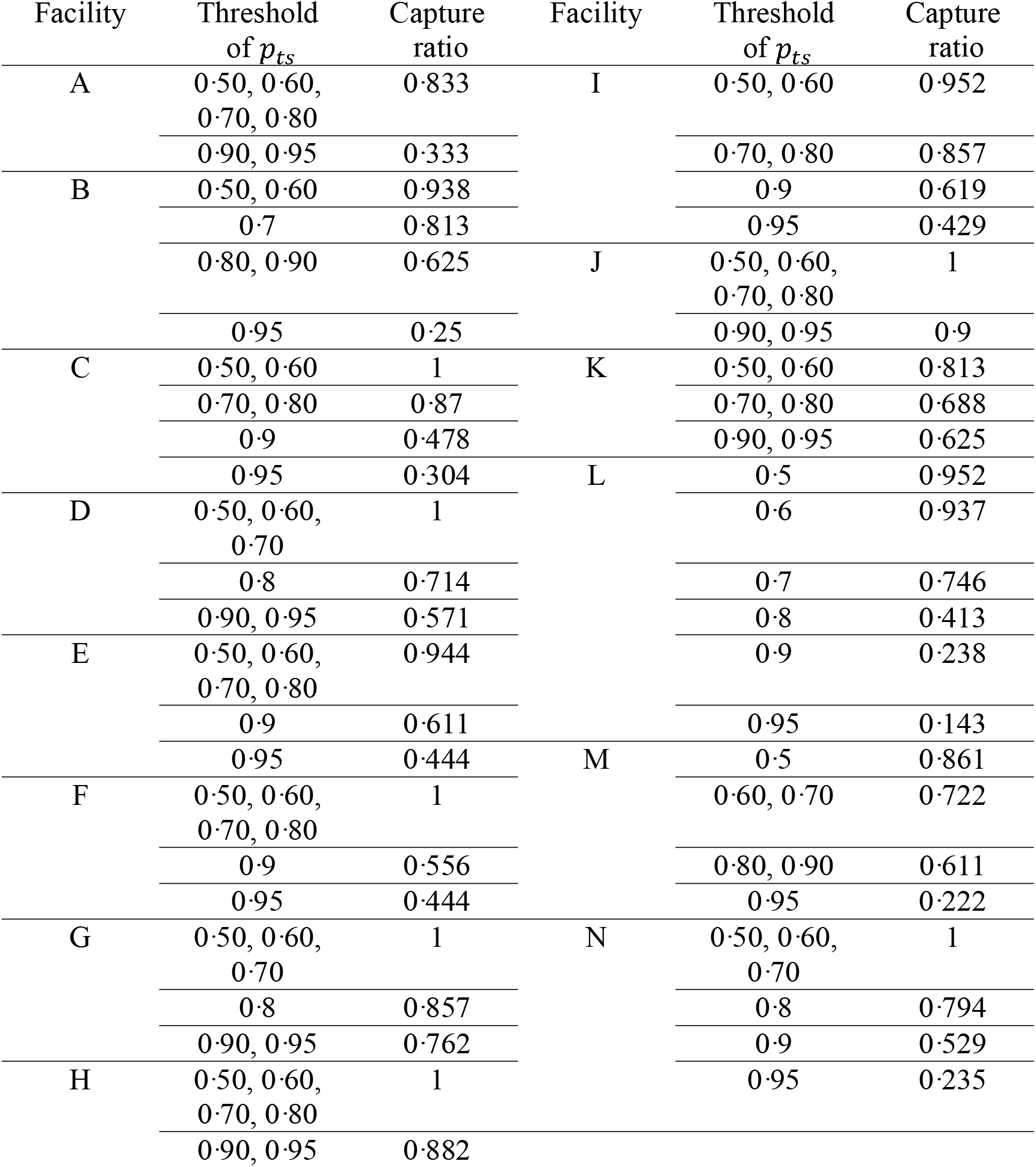
Sensitivity analysis per facility indicative of wastewater surveillance in considering the presence of incarcerated people with COVID-19 positive clinical tests at each facility from *p*_*ts*_ 0·50 to 0·95 and associated capture ratio.

The ideal thresholds are not uniform and need to be determined based on the results for each facility. Using an example of impact of a lower threshold at two sites, allows us to open up the discussion and in an efficient manner. Figure 2 provides a direct comparison between facility G, which demonstrates greater stability in response to *p*_*ts*_ thresholds, and facility L, which displays greater sensitivity to *p*_*ts*_ values when selecting *p*_*ts*_ thresholds of 0·8 and 0·5.

**Figure 2:**
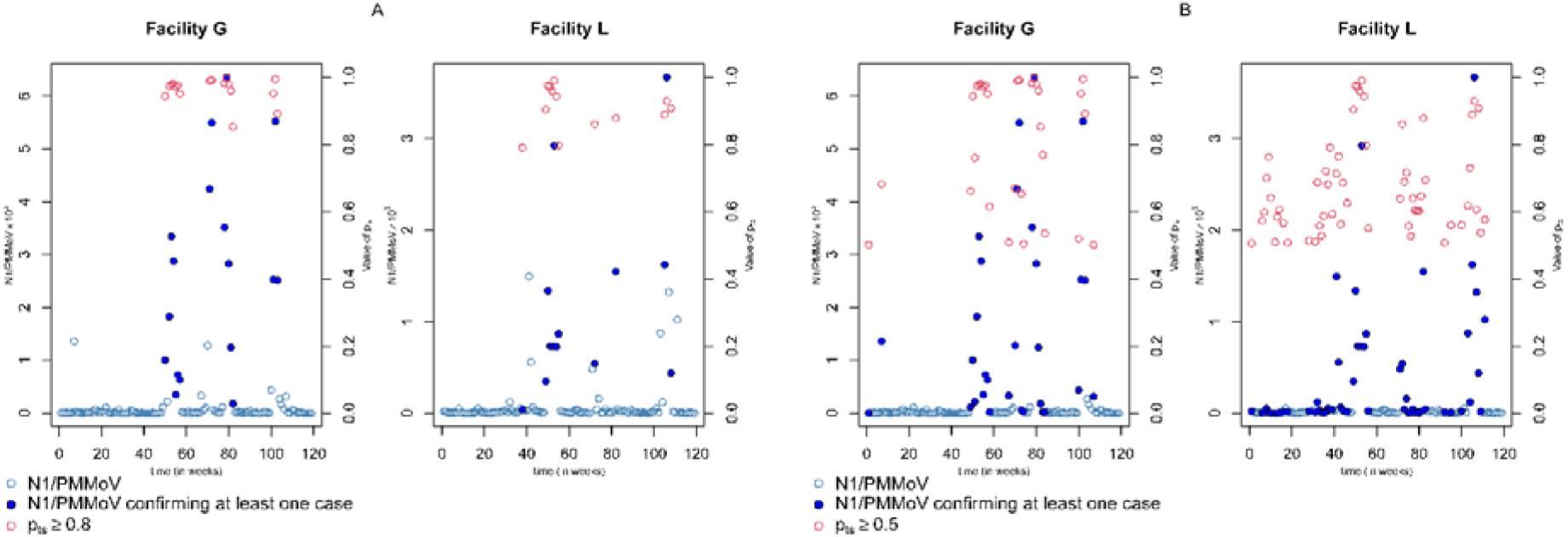
Testing impact of a lower threshold at two facilities. Temporal and spatial variability of SARS-CoV-2 (N1) copies per ml normalized by pepper mild mottle virus (PMMoV) copies per ml as a ratio compared of facility G and L for *p*_*ts*_ threshold 0·8 (A) versus 0·5 (B) of at least one positive COVID-19 case of an incarcerated person. Light blue outline circle indicates the wastewater SARS-CoV-2 (N1) copies per ml normalized by pepper mild mottle virus (PMMoV) copies per ml concentration ratio with no associated clinical case. Solid dark blue circle indicates the wastewater ratio associated with at least one clinical case with *p*_*ts*_ ≥ 0·8 (A) or 0·5 (B). The corresponding *p*_*ts*_ are plotted in red outline circle.

Ultimately, the determination of a *p*_*ts*_ threshold should involve a thoughtful examination of local epidemiological factors, which might include staff testing data, the presence of multiple viral variant strains, the quality of data influenced by sampling strategies and wastewater laboratory detection limits, testing capacity, the availability of healthcare resources, and prevailing public health policies. Regular monitoring and adjustment of this threshold may be necessary to adapt to changing conditions.

Other work has found the building-level detection threshold of SARS-CoV-2 in wastewater down to 1 true positive case is possible.^20^ But previous wastewater models have focused on estimating the numbers of infected individuals from a wastewater RNA concentration^6,13,21^; where our model differs rather is in estimating at least one individual from a wastewater RNA concentrations to guide a facility population health response regardless of the number of cases. Because we know conditions of confinement were a concern during the COVID-19 pandemic^22,23^ in terms of physical distancing and access to water, sanitation and hygiene facilities, it was additionally important but also unique for our model to consider the occupancy rate directly tied to both the clinical and wastewater concentrations. Wastewater surveillance can be used for rapid identification of infectious diseases in prisons,^24^ and our model provides a provides an additional tool that can be used to inform policy decisions.

## 4. Limitations

There are a number of other challenging factors which will require further exploration. These include accounting for the role of transient contributions to the wastewater which could include staff, visitors, and vendors. Additionally, we note the importance of having records of transfers of confirmed cases which was not practical to obtain during this public health emergency. While this study attempted to normalize SARS-CoV-2 concentrations for a contributing population and adjust for wastewater dilution using PMMOV concentrations, the use of alternative normalization factors such as wastewater flow rate paired to the sample collection date could also be useful. Future implementations of this type of environmental surveillance coupled with the rigorous statistical analysis as conducted herein, could consider evaluating the efficacy of different meta data.

## 5. Conclusion

Correctional facilities offer a unique environment for the application of wastewater virus detection as a tool to reduce population health risk. Although reports describing the use of longitudinal wastewater surveillance at corrections facilities across a state are sparse, our model has practical application for health equity. An essential application of the model in this study is the development of a model-derived criteria based on wastewater virus levels for early warning purposes as well as to better inform periods of declining population infection. In order to support ongoing real-time risk assessment, we modeled the SARS-CoV-2 (N1)/PMMoV ratio threshold associated with an 80% probability of at least one positive case being present. This probability value was used to illustrate one way to implement such a model and it is worth further exploration of whether reporting should include different levels of confidence or connections to different numbers of predicted cases. By assessing the consistency or discrepancy between these measures, policymakers can evaluate the effectiveness of existing strategies and determine if additional clinical testing measures should be proactively employed for the health of incarcerated populations.

## Data Availability

The computer code that implemented our model-based analysis will be made available immediately after publication. 

## Acknowledgements

This research was funded by the Commonwealth of Kentucky Department of Corrections Contracts PON2 527 2200001913 1 and PON2 527 2100001185. We would like to thank the Kentucky Department of Health, the Kentucky Department of Corrections, Eurofins Genomics LLC, and AECom for wastewater sample collection and analysis and feedback on the design and execution of this project. We thank Megan Beth Cushing for her efforts on an earlier version of this project.

## Contributors

KC and TS designed the wastewater testing and data collection approach. DH developed the mathematical model and designed the MCMC algorithm and PL generated the code and analyzed the data. DH and RHH drafted the first version of the manuscript with input from all authors. All authors contributed to the critical revision of the manuscript for important intellectual content. All authors have seen and approved the final version and agreed to its publication. DH and PL had full access to all the data in the study and take responsibility for the accuracy of the mathematical analysis.

