## Supplemental Material for "Projections of wastewater as an indicator of COVID-19 cases in corrections facilities: a modelling study"

### Appendix A. Summary of Facilities

Table A.1: Facility ID, operational capacity, and average inmate population (throughout study period).

| Facility | Occupational Capacity | Population Summary | | |
|---|---|---|---|---|
| | | Min | Max | Weekly Average |
| A | 300 | 119 | 265 | 170·09 |
| B | 320 | 195 | 301 | 227·47 |
| C | 1930 | 1374 | 1800 | 1604·34 |
| D | 982 | 389 | 804 | 485·61 |
| E | 733 | 485 | 673 | 552·17 |
| F | 914 | 402 | 776 | 484·10 |
| G | 1055 | 653 | 843 | 765·22 |
| H | 866 | 665 | 822 | 754·18 |
| I | 1062 | 789 | 976 | 890·93 |
| J | 1204 | 973 | 1068 | 1022·72 |
| K | 1270 | 989 | 1225 | 1138·09 |
| L | 1238 | 681 | 1119 | 952·90 |
| M | 621 | 193 | 572 | 409·07 |
| N | 730 | 392 | 693 | 497·96 |

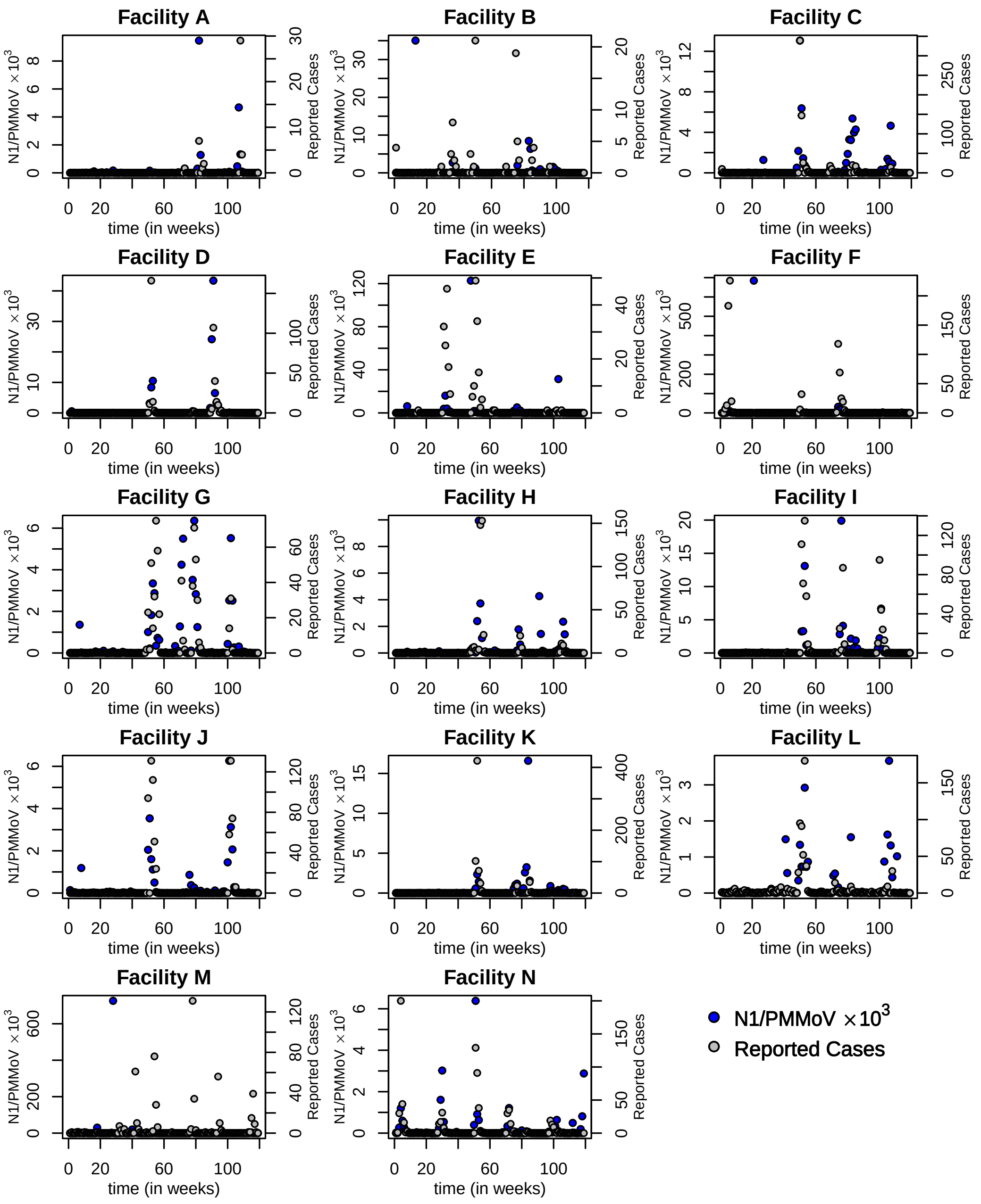

Figure A.1: The SARS-CoV-2 (N1) copies per ml normalized by pepper mild mottle virus (PMMoV) copies per ml as a ratio for each facility (right $y$-axis) throughout time compared to the observed number of inmate positive COVID-19 cases (left $y$-axis).

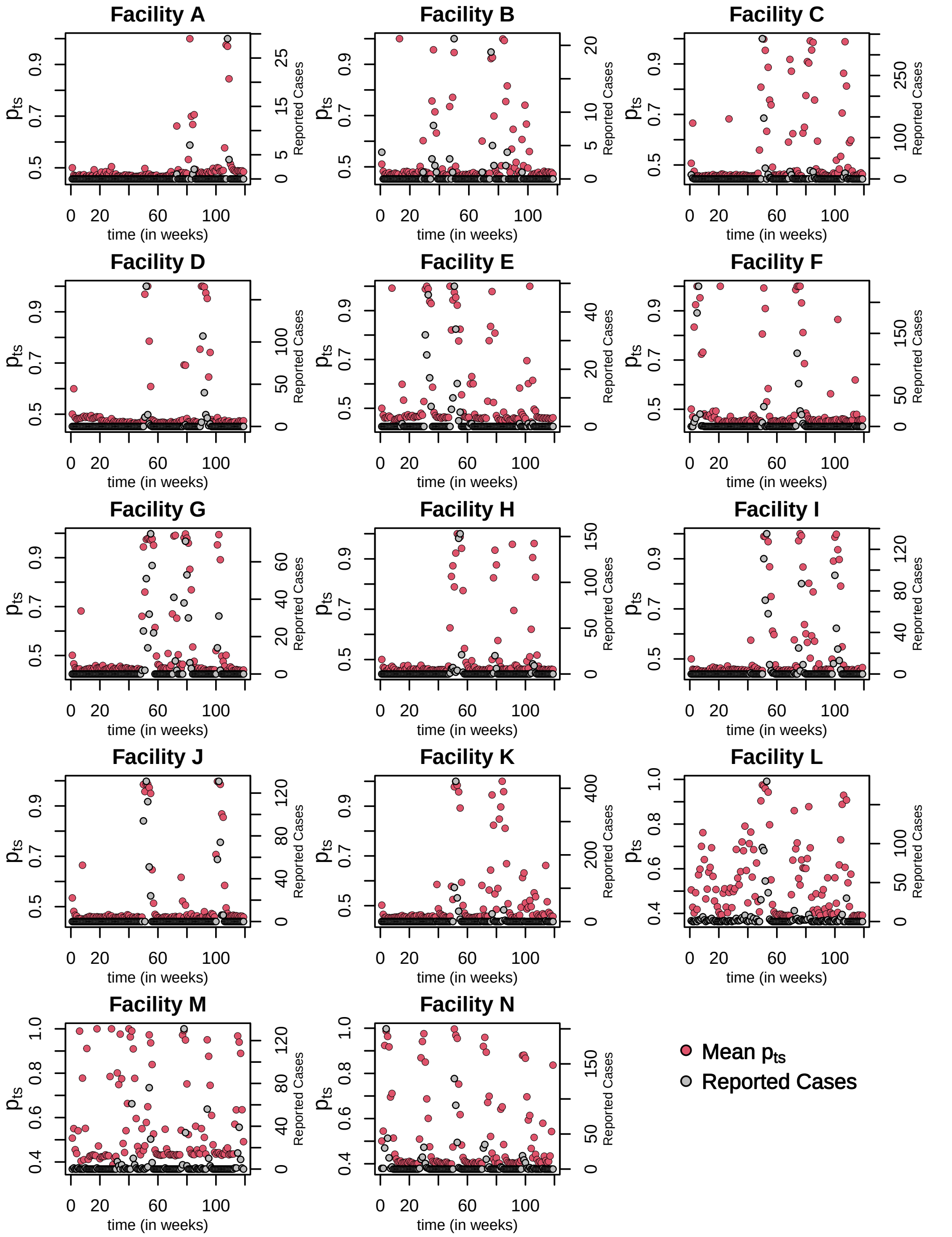

Figure A.2: Estimated $p_{ts}$ probabilities, the probability of at least one positive case versus the number of reported cases, $Z_{ts}$, by facility.

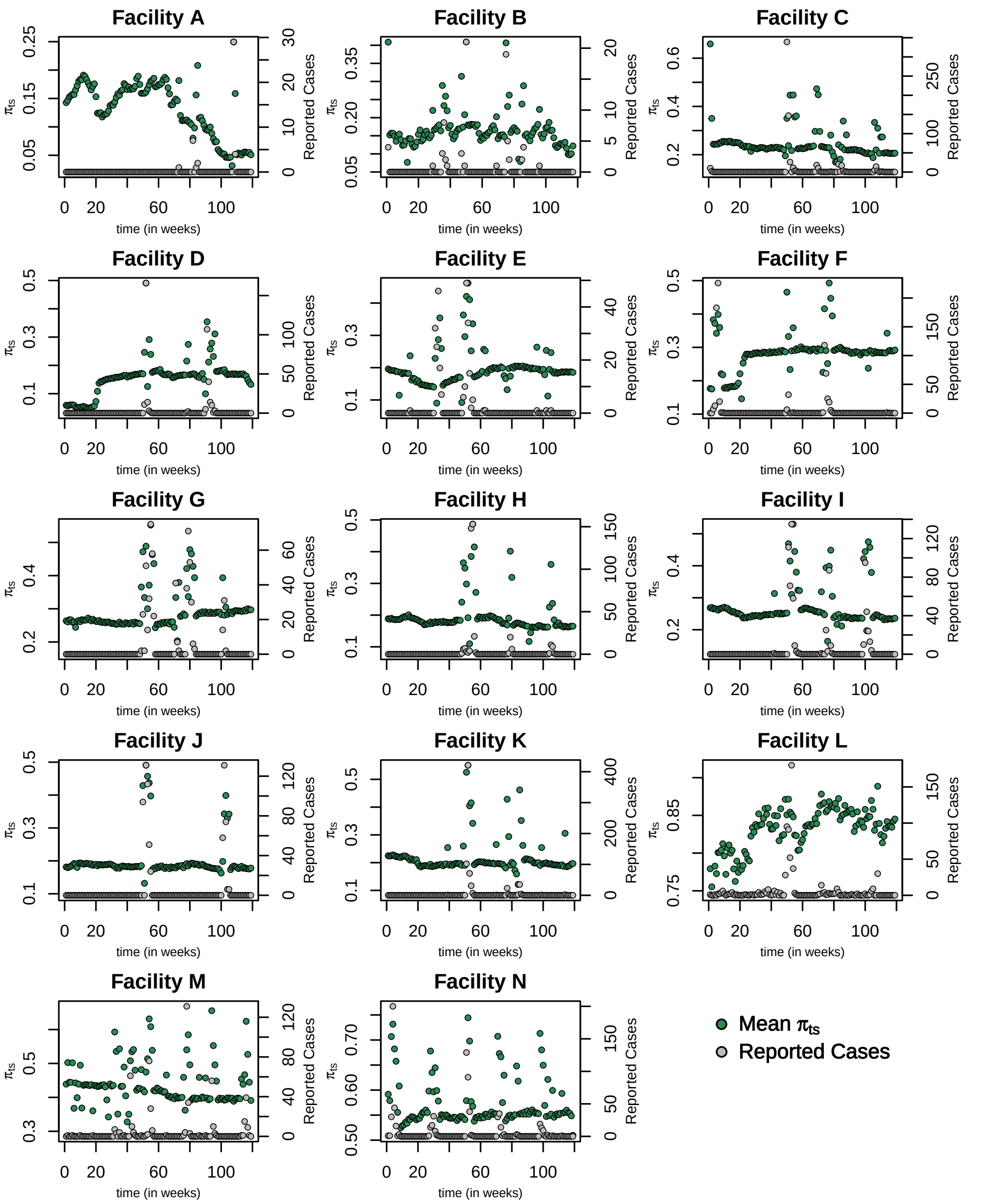

Figure A.3: Estimated reporting probabilities $\pi_{ts}$ for each facility $s$ compared to the reported number of positive inmate COVID-19 cases.

### Appendix B. Hierarchical Spatial-temporal Model with Lags for Underreporting Data

Let $N_{ts}$ and $Z_{ts}$ denote, respectively, the true but unobserved number of positive COVID-19 cases and the recorded (observed) number of cases for location $s \in \{1, 2, \cdots, J\}$ at time $t$, $t = 1, 2, 3, \cdots, N$. Assume each location $s$, $s \in \{1, 2, 3, \cdots, J\}$ has different independent reporting probabilities $\pi_{ts}$ at time $t$ due to the distinct situations present in different correctional facilities such as sampling methods and testing capabilities. Then conditional on the true positive cases number $N_{ts}$, the time series of counts $Z_{ts}$ reported at time $t$ at location $s$ follows a Binomial distribution with probability $\pi_{ts}$:

$$Z_{ts}|N_{ts} \sim Binomial\left(N_{ts}, \pi_{ts}\right) \tag{B.1}$$

We will assume that the reporting probabilities $\pi_{ts}$ in (B.1) depend on a set of covariates such as inmate population and capacity ratio. Denote the design matrix containing these covariates by $X_{ts}^{(1)}$ and the reporting probability $\pi_{ts}$ is modeled by a logistic regression on $X_{ts}^{1}$:

$$\log\left(\frac{\pi_{ts}}{1-\pi_{ts}}\right) = X_{ts}^{(1)}\vec{\beta_s} + \varepsilon_{ts} = \psi_{ts} \tag{B.2}$$

where $\vec{\beta_s}$ is the $N \times 1$ vector of the regression coefficients, $\vec{\beta_s}$ has normal prior with mean $m_{\beta_0}$ and covariance matrix $P_{\beta_0}$. The white noise error term $\varepsilon_{ts}$ has the normal prior with mean $\mu_{\varepsilon_{0,s}}$ and variance $\sigma^2_{\varepsilon_{0,s}}$.

Assume the number of true positive COVID-19 cases $N_{ts}$ at location $s$ at time $t$ follows a latent Poisson distribution with rate $\lambda_{ts}$, which is a Gamma distribution with shape parameter $h = 1$ and scale parameter $\frac{p_{ts}}{1-p_{ts}}$.

$$N_{ts}|\lambda_{ts} \sim Poisson\left(\lambda_{ts}\right) \tag{B.3}$$

$$\lambda_{ts}|p_{ts} \sim \mathcal{G}amma\left(h, \frac{p_{ts}}{1-p_{ts}}\right) \tag{B.4}$$

If we marginalize over $\lambda_{ts}$, we get a negative binomial marginal distribution for $N_{ts}$:

$$P(N_{ts} = k \,|\, h, p_{ts}) = \frac{\Gamma(k+h)}{k!\Gamma(h)}(1-p_{ts})^h p_{ts}^{N_{ts}} \tag{B.5}$$

Thus $p_{ts}$ is the probability of at least one positive case at $t$ in the facility $s$, i.e, $p_{ts} = P(N_{ts} \geq 1)$. To capture the hidden dynamic lagged effects, $p_{ts}$ is modeled as a logistic regression embedded with an auto-regressive latent process $\gamma_{ts}$ with order 1 (AR(1) process). The MCMC algorithm for the more generalized higher-order AR(p) can be extended easily based on the current model.

$$\log\left(\frac{p_{ts}}{1-p_{ts}}\right) = X_{ts}^{(2)} + \gamma_{ts} = \phi_{ts} \tag{B.6}$$

$$\gamma_{ts} = a_s \gamma_{t-1,s} + e_{ts} \tag{B.7}$$

where $X_{ts}^{(2)}$ is a matrix of relevant factors related to true positive cases. In this paper, we use the normalized SARS-CoV-2 (N1) copies per ml normalized by pepper mild mottle virus (PMMoV) copies per ml as a ratio as $X_{ts}^{(2)}$. And the error term $e_{ts}$ follows normal distribution with mean 0 and variance $\sigma^2_{e0,s}$. The parameter $a_s$ has truncated normal distribution on $(-1, 1)$ as the prior, that is $a_s \sim Normal\left(\mu_{a0,s}, \sigma^2_{a0,s}\right)\mathcal{I}(-1 < a_s < 1)$.

In the realm of spatial-temporal models with lagged effects, it is observed that the majority of models do not take into account the underreporting situation from the systematic review of COVID-19 models[1] and recent various publications.[2–6] This highlights a critical gap in the existing literature. Moreover, current underreport count time series models, such as the widely used Poisson-Logistic (Pogit) model,[7] do not incorporate lagged effects. This limitation restricts their ability to capture the temporal dynamics and delays in reporting COVID-19 cases accurately.

In addition, existing hierarchical count models for the under-reporting type data often rely on benchmarks like mortality rates[8] or overall population numbers from census data[9] to infer the true number of positive cases. However, when studying the relationship between positive COVID-19 cases and wastewater virus concentration, the dynamic movement of the population presents a significant challenge. The movement of individuals within a community, encompassing their residences, workplaces, and public spaces, introduces complexities in accurately tracking the corresponding population numbers within specific sewer sheds. This dynamic nature of population movement makes it difficult to establish a direct and precise correlation between the number of positive cases and the virus concentration in wastewater samples from specific sewer sheds.

In the context of studying prison cases, these issues can be circumvented. The controlled environment of correctional facilities allows for more accurate tracking of the population and their movement, minimizing the challenges associated with population dynamics. By focusing on corrections facility cases, the study can provide valuable insights into the relationship between positive COVID-19 cases and wastewater virus concentration, avoiding the complexities arising from population movement in the broader community.

### Appendix C. MCMC sampling

If we marginalize over $\lambda_{ts}$, we obtain a negative binomial distribution for $N_{ts}$,

$$p\left(N_{ts}|h, p_{ts}\right) \propto \left(1 - p_{ts}\right)^h \, p_{ts}^{N_{ts}} \ , \tag{C.1}$$

ignoring constant of proportionality, i.e., $N_{ts}|h, p_{ts} \sim \text{NegBin}\left(h, p_{ts}\right)$. Using a logit transform, we obtain

$$p_{ts} = \frac{e^{\phi_{ts}}}{1 + e^{\phi_{ts}}} \Rightarrow \quad p\left(N_{ts}|h, p_{ts}\right) \propto \frac{\left(e^{\phi_{ts}}\right)^{N_{ts}}}{\left(1 + e^{\phi_{ts}}\right)^{h + N_{ts}}} \tag{C.2}$$

In this study, we employ a Pólya-Gamma data-augmentation strategy to facilitate the fast fully Bayesian inference using Gibbs sampling. We make use of the following theorem studied by Nicholas Polson and his coauthors:[10]

**Theorem 1.** *Let $p(\omega)$ denote the density of the random variable $\omega \sim \mathcal{PG}(b, 0)$ for $b > 0$, where $\mathcal{PG}(\cdot, \cdot)$ denotes the Polya-Gamma distribution. Then, the following integral identity holds for all $a \in \mathbb{R}$,*

$$\frac{\left(e^{\psi}\right)^a}{\left(1 + e^{\psi}\right)^b} = 2^{-b} e^{\kappa \psi} \int_0^{\infty} e^{\omega \, \psi^2/2} p(\omega) d\omega \tag{C.3}$$

*where $\kappa = a - b/2$. Moreover, treating the integrand in (C.3) as an unnormalized joint density $(\psi|\omega)$ gives rise to the conditional distribution*

$$p(\omega|\psi) = \frac{e^{-\omega\psi^2/2} \, p(\omega)}{\int_0^{\infty} e^{-\omega\psi^2/2} \, p(\omega) \, d\omega} \ , \tag{C.4}$$

*which is also in the Polya-Gamma class: $(\omega|\psi) \sim \mathcal{PG}\left(b, \psi\right)$.*

**Sampling of $a_s$:**

Let $\vec{\gamma_s} = [\gamma_{t-1,s}]_{t=1}^{n}$. We find the conditional posterior distribution of $a_s$,

$$p\left(a_s\,|\,\cdot\right) \propto \prod_{t=1}^{n} p\left(N_{ts}\,|\,\phi_{ts}\right) p\left(\phi_{ts}\,|\,a_s, \sigma^2_{e_{0,s}}\right) p(a_s) \tag{C.5}$$

$$\propto \prod_{t=1}^{n} \frac{\left(e^{\phi_{ts}}\right)^{N_{ts}}}{\left(1+e^{\phi_{ts}}\right)^{h+N_{ts}}} \frac{e^{\frac{1}{2\sigma^2_{e_{0,s}}}(\phi_{ts}-X^{(2)}_{ts}-a_s\vec{\gamma_s})^2}}{\sqrt{2\pi\sigma^2_{e_{0,s}}}} \frac{e^{\frac{1}{2\sigma^2_{a_{0,s}}}(a_s-\mu_{a_{0,s}})^2}}{\sqrt{2\pi\sigma^2_{a_{0,s}}}} \tag{C.6}$$

Using the integral identity (C.3), we can re-write the above in terms of $\phi_{ts}$,

$$p\left(a_s\,|\,\cdot\right) \propto \prod_{t=1}^{n} 2^{-N_{ts}-h} e^{\kappa^{(2)}_{ts}\phi_{ts}} \int_0^{\infty} e^{\omega^{(2)}_{ts}\phi^2_{ts}/2}\, p\left(\omega^{(2)}\right) d\omega^{(2)} \cdot \frac{e^{\frac{1}{2\sigma^2_{e_{0,s}}}(\phi_{ts}-X^2_{ts}-a_s\vec{\gamma_s})^2}}{\sqrt{2\pi\sigma^2_{e_{0,s}}}} \frac{e^{\frac{1}{2\sigma^2_{a_{0,s}}}(a_s-\mu_{a_{0,s}})^2}}{\sqrt{2\pi\sigma^2_{a_{0,s}}}} \tag{C.7}$$

where $\kappa^{(2)}_{ts} = N_{ts} - \frac{h+N_{ts}}{2} = \frac{N_{ts}-h}{2}$. It follows from Theorem 1 that the mixing distribution is $\omega^{(2)}_{ts}|\phi_{ts} \sim \mathcal{PG}\left(N_{ts}+h, \phi_{ts}\right)$. Let $\vec{\kappa_s}^{(2)} = \left(\left[\kappa^{(2)}_{ts}\right]_{t=1}^{n}\right)^T$, $Z^{(2)}_s = \left(\left[\frac{ks^{(2)}}{\omega^{(2)}_{ts}}\right]_{t=1}^{n}\right)^T$ and $\Omega^{(2)}_s = \text{diag}\left(\omega^{(2)}_{1s}, \cdots, \omega^{(2)}_{ns}\right)$. Then,

$$p\left(a_s\,|\,\cdot\right) \propto \int_0^{\infty} \prod_{t=1}^{n} e^{-\frac{\omega^{(2)}_{ts}}{2}\left(\frac{\kappa^{(2)}_{ts}}{\omega^{(2)}_{ts}} - \phi^2_{ts}\right)^2} p\left(\omega^{(2)}\right) d\omega^{(2)} \cdot \frac{e^{\frac{1}{2\sigma^2_{e_{0,s}}}(\phi_{ts}-X^2_{ts}-a_s\vec{\gamma_s})^2}}{\sqrt{2\pi\sigma^2_{e_{0,s}}}} \frac{e^{\frac{1}{2\sigma^2_{a_{0,s}}}(a_s-\mu_{a_{0,s}})^2}}{\sqrt{2\pi\sigma^2_{a_{0,s}}}} \tag{C.8}$$

$$\propto \int_0^{\infty} e^{-(Z^{(2)}_s-X^{(2)}_s-a_s\vec{\gamma_s}-\vec{e_s})^T\Omega^{(2)}_s(Z^{(2)}_s-X^{(2)}_s-a_s\vec{\gamma_s}-\vec{e_s})/2} p(w^{(2)}) d\omega^{(2)} \tag{C.9}$$
$$\cdot\, e^{(\phi_s-X^{(2)}_s-a_s\vec{\gamma_s})^T(\sigma_{e_{0,s}})^{-1}\mathbb{I}(\phi_s-X^{(2)}_s-a_s\vec{\gamma_s})} e^{\frac{1}{2\sigma^2_{a_{0,s}}}(a_s-\mu_{a_{0,s}})^2} \mathcal{I}(-1<a_s<1)$$

Hence, the conditional posterior distribution of $a_s$ is,

$$a_s \sim Normal\left(\mu_{a_s}, V_{a_s}\right)\mathcal{I}(-1 < a_s < 1) \quad , \tag{C.10}$$

$$\text{where} \quad V_{a_s} = \left[\vec{\gamma_s}^T \Omega_s^{(2)} \vec{\gamma_s} + \vec{\gamma_s}^T (\sigma^2_{e_{0,s}})^{-1} \mathbb{I}\ \vec{\gamma_s} + (\sigma^2_{a_{0,s}})^{-1}\right]^{-1} \quad , \tag{C.11}$$

$$\text{and} \quad \mu_{a_s} = V_{a_s}\left[\vec{\gamma_s}^T \Omega_s^{(2)}\left(Z_s^{(2)} - X_s^{(2)} - \vec{e_s}\right) + \vec{\gamma_s}^T (\sigma^2_{e_{0,s}})^{-1}\mathbb{I}\left(\phi_s - X_s^{(2)}\right) + (\sigma^2_{a_{0,s}})^{-1}\mu_{a_{0,s}}\right] \tag{C.12}$$

**Sampling of $\vec{e_s}$:**
Let $\vec{e_s} = \left([e_{ts}]_{t=1}^{n}\right)^T$. Using the same derivation as above and equation (C.9), we find the distribution of $\vec{e_s}$,

$$p\left(\vec{e_s} \mid \cdot\right) \propto \prod_{t=1}^{n} p\left(N_{ts} \mid \phi_{ts}\right) p\left(e_{ts}\right) \tag{C.13}$$

$$\propto \int_0^\infty e^{-(Z_s^{(2)} - X_s^{(2)} - a_s\vec{\gamma_s} - \vec{e_s})^T \Omega_s^{(2)} (Z_s^{(2)} - X_s^{(2)} - a_s\vec{\gamma_s} - \vec{e_s})/2}\ p(\omega^{(2)}) d\omega^{(2)} \cdot \frac{e^{\vec{e_s}^T (\sigma^2_{e_{0,s}})^{-1}\mathbb{I}\vec{e_s}}}{\sqrt{2\pi\sigma^2_{e_{0,s}}}} \tag{C.14}$$

Thus, the distribution of $\vec{e_s}$ is,

$$\vec{e_s} \sim Normal\left(\mu_{e_s}, V_{e_s}\right) \quad , \tag{C.15}$$

$$\text{where} \quad V_{e_s} = \left[\Omega_s^{(2)} + (\sigma^2_{e_{0,s}})^{-1}\mathbb{I}\right]^{-1} \tag{C.16}$$

$$\text{and} \quad \mu_{e_s} = V_{e_s}\left[\Omega_s^{(2)}\left(Z_s^{(2)} - X_s^{(2)} - a_s\vec{\gamma_s}\right)\right] \tag{C.17}$$

**Sampling of $\vec{\beta_s}$:**
To find the conditional posterior distribution of $\vec{\beta_s}$, we look at the likelihood

function

$$p\left(\vec{\beta}_s \,|\, \cdot\right) \propto \prod_{t=1}^{n} p\left(Z_{ts} \,|\, N_{ts}, \psi_{ts}\right) p\left(\psi_{ts} \,|\, \varepsilon_{ts}\right) p\left(\vec{\beta}_s\right) \tag{C.18}$$

$$\propto \prod_{t=1}^{n} \binom{N_{ts}}{Z_{ts}} \left(\frac{e^{\psi_{ts}}}{1+e^{\psi_{ts}}}\right)^{Z_{ts}} \left(\frac{1}{1+e^{\psi_{ts}}}\right)^{N_{ts}-Z_{ts}} \cdot p\left(\psi_{ts} \,|\, \varepsilon_{ts}\right) p\left(\vec{\beta}_s\right) \tag{C.19}$$

$$\propto \prod_{t=1}^{n} \binom{N_{ts}}{Z_{ts}} \frac{\left(e^{\psi_{ts}}\right)^{Z_{ts}}}{\left(1+e^{\psi_{ts}}\right)^{N_{ts}}} \frac{e^{\frac{-1}{2\sigma^2_{\varepsilon_{0,s}}}(\psi_{ts}-X^{(1)}_{ts}\vec{\beta}_s-\mu_{\varepsilon_{0,s}})^2}}{\sqrt{2\pi\sigma^2_{\varepsilon_{0,s}}}} \cdot e^{-(\vec{\beta}_s-m_{\beta_0})^T P^{-1}_{\beta_0}(\vec{\beta}_s-m_{\beta_0})/2} \tag{C.20}$$

Applying integral identity (C.3), we can write the likelihood above in terms of $\psi_{ts}$:

$$p\left(\vec{\beta}_s \,|\, \cdot\right) \propto \prod_{t=1}^{n} 2^{-N_{ts}} e^{\kappa^{(1)}_{ts}\psi_{ts}} \int_0^\infty e^{\omega^{(1)}_{ts}\psi^2_{ts}/2} p\left(\omega^{(1)}\right) d\omega^{(1)} \cdot \frac{e^{\frac{-1}{2\sigma^2_{\varepsilon_{0,s}}}(\psi_{ts}-X^{(1)}_{ts}\vec{\beta}_s-\mu_{\varepsilon_{0,s}})^2}}{\sqrt{2\pi\sigma^2_{\varepsilon_{0,s}}}} e^{-(\vec{\beta}_s-m_{\beta_0})^T P^{-1}_{\beta_0}(\vec{\beta}_s-m_{\beta_0})/2} , \tag{C.21}$$

where $\kappa^{(1)}_{ts} = Z_{ts} - N_{ts}/2$. Thus, Theorem 1 implies that the mixing distribution is $\omega^{(2)}_{ts}|\phi_{ts} \sim \mathcal{PG}\left(N_{ts}, \psi_{ts}\right)$.

Let $\vec{\kappa_s}^{(1)} = \left(\left[\kappa_{ts}^{(1)}\right]_{t=1}^{n}\right)^T$, $Z_s^{(1)} = \left(\left[\frac{ks^{(1)}}{\omega_{ts}^{(1)}}\right]_{t=1}^{n}\right)^T$ and $\Omega_s^{(1)} = \text{diag}\left(\omega_{1s}^{(1)}, \cdots, \omega_{ns}^{(1)}\right)$. Then,

$$p\left(\vec{\beta_s} \,|\, \cdot\right) \propto \int_0^\infty \prod_{t=1}^{n} e^{-\frac{\omega_{ts}^{(1)}}{2}\left(\frac{\kappa_{ts}^{(1)}}{\omega_{ts}^{(1)}} - \psi_{ts}^2\right)^2} p\left(\omega^{(1)}\right) d\omega^{(1)}$$

$$\cdot\, e^{\frac{-1}{2\sigma^2_{\varepsilon_{0,s}}}(\psi_{ts} - X_{ts}^{(1)}\vec{\beta_s} - \mu_{\varepsilon_{0,s}})^2} e^{-(\vec{\beta_s} - m_{\beta_0})^T P_{\beta_0}^{-1}(\vec{\beta_s} - m_{\beta_0})/2} \tag{C.22}$$

$$\propto \int_0^\infty e^{-\frac{1}{2}\left(Z_s^{(1)} - X_s^{(1)}\vec{\beta_s} - \vec{\varepsilon_s}\right)^T \Omega_s^{(1)}\left(Z_s^{(1)} - X_s^{(1)}\vec{\beta_s} - \vec{\varepsilon_s}\right)} p\left(\omega^{(1)}\right) d\omega^{(1)} \tag{C.23}$$

$$\cdot\, e^{-\frac{1}{2}\left(\psi_s - X_s^{(1)}\vec{\beta_s} - \mu_{\varepsilon_{0,s}}\right)^T (\sigma^2_{\varepsilon_{0,s}})^{-1}\mathbb{I}\left(\psi_s - X_s^{(1)}\vec{\beta_s} - \mu_{\varepsilon_{0,s}}\right)}$$

$$\cdot\, e^{-\frac{1}{2}\left(\vec{\beta_s} - m_{\beta_0}\right)^T P_{\beta_0}^{-1}\left(\vec{\beta_s} - m_{\beta_0}\right)}$$

Completing the square yields the following distribution for $\vec{\beta_s}$:

$$\vec{\beta_s} \sim \; Normal\left(\mu_{\vec{\beta_s}}, V_{\vec{\beta_s}}\right) \tag{C.24}$$

$$\text{where} \quad V_{\vec{\beta_s}} = \left[X_s^{(1)}\left(\Omega_s^{(1)} + (\sigma^2_{\varepsilon_{0,s}})^{-1}\mathbb{I}\right) + P_{\beta_0}^{-1}\right]^{-1} \text{and} \tag{C.25}$$

$$\mu_{\vec{\beta_s}} = V_{\vec{\beta_s}}\left[X_s^{(1)}\Omega_s^{(1)}\left(Z_s^{(1)} - \vec{\varepsilon_s}\right) + (\sigma^2_{\varepsilon_{0,s}})^{-1}X_s^{(1)}\left(\psi_s - \mu_{\varepsilon_{0,s}}\right) + P_{\beta_0}^{-1}m_{\beta_0}\right]. \tag{C.26}$$

**Sampling of $\varepsilon_{ts}$:**
Let $\vec{\varepsilon_s} = \left([\varepsilon_{ts}]_{t=1}^{n}\right)^T$. Using the same decomposition shown in equation (C.23), the joint likelihood for $\vec{\varepsilon_s}$ is

$$p\left(\vec{\varepsilon_s} \,|\, \cdot\right) \propto \prod_{t=1}^{n} p\left(Z_{ts} \,|\, N_{ts}, \psi_{ts}\right) p\left(\vec{\varepsilon_s}\right) \tag{C.27}$$

$$\propto \int_0^\infty e^{-\frac{1}{2}\left(Z_s^{(1)} - X_s^{(1)}\vec{\beta_s} - \vec{\varepsilon_s}\right)^T \Omega_s^{(1)}\left(Z_s^{(1)} - X_s^{(1)}\vec{\beta_s} - \vec{\varepsilon_s}\right)} p\left(\omega^{(1)}\right) d\omega^{(1)}$$

$$\cdot\, e^{-\frac{1}{2}\left(\vec{\varepsilon_s} - \mu_{\varepsilon_{0,s}}\right)^T (\sigma^2_{\varepsilon_{0,s}})^{-1}\mathbb{I}\left(\vec{\varepsilon_s} - \mu_{\varepsilon_{0,s}}\right)} \;. \tag{C.28}$$

Thus, $\vec{\varepsilon_s}$ follows a normal distribution with parameters,

$$\vec{\varepsilon_s} \sim Normal\left(\mu_{\vec{\varepsilon_s}}, V_{\vec{\varepsilon_s}}\right) \tag{C.29}$$

$$\text{where} \quad V_{\vec{\varepsilon_s}} = \left[\Omega_s^{(1)} + \left(\sigma^2_{\varepsilon_{0,s}}\right)^{-1} \mathbb{I}\right]^{-1} \tag{C.30}$$

$$\text{and} \quad \mu_{\vec{\varepsilon_s}} = \left[\Omega_s^{(1)} \left(Z_s^{(1)} - X_s^{(1)} \vec{\beta_s}\right) + \left(\sigma^2_{\varepsilon_{0,s}}\right)^{-1} \mu_{\varepsilon_{0,s}}\right] \cdot \tag{C.31}$$

**Sampling of $\lambda_{ts}$:**
To sample $\lambda_{ts}$, we use the negative binomial distribution of $N_{ts}$,

$$p\left(\lambda_{ts} \mid \cdot\right) \propto p\left(N_{ts} \mid \lambda_{ts}\right) p\left(\lambda_{ts}\right) \tag{C.32}$$

$$\propto \frac{\left(\lambda_{ts}\right)^{N_{ts}} e^{-\lambda_{ts}}}{N_{ts}!} \cdot \frac{1}{\Gamma(h)\left(\frac{p_{ts}}{1-p_{ts}}\right)^{h}} \left(\lambda_{ts}\right)^{h-1} e^{-\lambda_{ts}/\left(\frac{p_{ts}}{1-p_{ts}}\right)} \tag{C.33}$$

$$\propto \left(\lambda_{ts}\right)^{N_{ts}+h-1} e^{-\lambda_{ts}\left(1+\frac{p_{ts}}{1-p_{ts}}\right)} \tag{C.34}$$

$$\propto \left(\lambda_{ts}\right)^{N_{ts}+h-1} e^{-\lambda_{ts}\left(1+\frac{1}{p_{ts}}-1\right)} \tag{C.35}$$

$$\propto \left(\lambda_{ts}\right)^{N_{ts}+h-1} e^{-\frac{\lambda_{ts}}{p_{ts}}} \tag{C.36}$$

Hence, $\lambda_{ts}$ follows a Gamma distribution with shape parameter $N_{ts} + h$ and scale parameter $p_{ts}$, i.e.,

$$\lambda_{ts} \sim \mathcal{G}amma\left(N_{ts} + h, p_{ts}\right) \tag{C.37}$$

**Sampling of $N_{ts}$:**
Given that $Z_{ts} \mid N_{ts} \sim Binomial\left(N_{ts}, \pi_{ts}\right)$ and $N_{ts} \sim Poisson\left(\lambda_{ts}\right)$, we have that the marginal distribution of $Z_{ts}$ follows a Poisson distribution with rate parameter $\pi_{ts}\lambda_{ts}$,

$$Z_{ts} \sim Poisson\left(\pi_{ts}\lambda_{ts}\right) \tag{C.38}$$

By Bayes rules, we have that

$$N_{ts} - Z_{ts} \sim Poisson\left(\left(1 - \pi_{ts}\right)\lambda_{ts}\right) \tag{C.39}$$

### Appendix D. Parameter Estimations

Table D.2: Estimated $\vec{\beta_s}$ coefficients with their 90% Confidence Interval for all locations.

| Facility | Coef | Mean | Median | 90% Conf. Int lower | 90% Conf. Int upper |
|---|---|---|---|---|---|
| A | $\beta_1$ | -1·98301 | -2·87585 | -4·4810 | 1·2757 |
| | $\beta_2$ | -0·0020204 | 0·003236 | -1·6456 | 1·6315 |
| B | $\beta_1$ | -1·30041 | -1·92976 | -3·1829 | 1·3736 |
| | $\beta_2$ | 0·11357 | 0·12165 | -1·5264 | 1·7255 |
| C | $\beta_1$ | -0·80471 | -1·23861 | -2·0319 | 1·2903 |
| | $\beta_2$ | 0·014857 | 0·021477 | -1·6388 | 1·6537 |
| D | $\beta_1$ | -2·06116 | -2·82419 | -4·7767 | 1·3085 |
| | $\beta_2$ | 0·026494 | 0·027866 | -1·5996 | 1·6605 |
| E | $\beta_1$ | -1·10048 | -1·67059 | -2·6604 | 1·3374 |
| | $\beta_2$ | 0·048162 | 0·037346 | -1·6061 | 1·7042 |
| F | $\beta_1$ | -0·94115 | -1·41497 | -2·5246 | 1·4503 |
| | $\beta_2$ | 0·16089 | 0·16539 | -1·4811 | 1·7988 |
| G | $\beta_1$ | -0·71879 | -1·05832 | -2·0139 | 1·3215 |
| | $\beta_2$ | 0·039300 | 0·036963 | -1·5996 | 1·6957 |
| H | $\beta_1$ | -0·98055 | -1·49157 | -2·3781 | 1·3095 |
| | $\beta_2$ | 0·0084821 | 0·016313 | -1·6058 | 1·6455 |
| I | $\beta_1$ | -0·70491 | -1·09014 | -1·8739 | 1·3001 |
| | $\beta_2$ | 0·022349 | 0·021148 | -1·6176 | 1·6657 |
| J | $\beta_1$ | -0·96685 | -1·45703 | -2·4120 | 1·3141 |

Table D.2: Estimated $\vec{\beta_s}$ coefficients with their 90% Confidence Interval for all locations.

| Facility | Coef | Mean | Median | 90% Conf. Int | |
|---|---|---|---|---|---|
| | | | | lower | upper |
| | $\beta_2$ | 0·036615 | 0·040357 | -1·6142 | 1·6813 |
| K | $\beta_1$ | -0·83426 | -1·28090 | -2·1207 | 1·3438 |
| | $\beta_2$ | 0·055580 | 0·046813 | -1·5663 | 1·6813 |
| L | $\beta_1$ | 1·26970 | 1·59546 | -1·3200 | 3·4638 |
| | $\beta_2$ | -0·005757 | -0·013282 | -1·6650 | 1·6647 |
| M | $\beta_1$ | -0·18888 | -0·30551 | -1·3055 | 1·3144 |
| | $\beta_2$ | 0·050945 | 0·056434 | -1·5930 | 1·6640 |
| N | $\beta_1$ | 0·32784 | 0·37297 | -1·1410 | 1·5896 |
| | $\beta_2$ | 0·13592 | 0·13555 | -1·4972 | 1·7790 |

Table D.3: Estimated ratios for $a_s$ by location along with the 90% confidence interval.

| Facility | Mean | Median | 90% Conf. Int | |
|---|---|---|---|---|
| | | | lower | upper |
| A | 0·064371 | 0·062732 | -0·104865 | 0·235365 |
| B | 0·075738 | 0·075819 | -0·091659 | 0·239276 |
| C | 0·119560 | 0·118169 | -0·024537 | 0·271463 |
| D | 0·058017 | 0·057467 | -0·105399 | 0·224653 |
| E | 0·249851 | 0·250210 | 0·103090 | 0·395005 |
| F | 0·122342 | 0·122619 | -0·034707 | 0·280635 |
| G | 0·318125 | 0·318466 | 0·184318 | 0·449081 |
| H | 0·279283 | 0·278859 | 0·141133 | 0·413762 |
| I | 0·178254 | 0·177557 | 0·025735 | 0·328457 |
| J | 0·303776 | 0·304538 | 0·169531 | 0·435167 |
| K | 0·306361 | 0·306897 | 0·187298 | 0·424489 |
| L | 0·334436 | 0·333574 | 0·216690 | 0·454484 |
| M | 0·274937 | 0·275520 | 0·137353 | 0·411050 |
| N | 0·416922 | 0·416648 | 0·299533 | 0·536780 |

### Appendix E. Summary of SARS-CoV-2 (N1)/PMMoV ratio with alternative probabilities

Table E.4: Selected SARS-CoV-2 (N1) copies per ml normalized by pepper mild mottle virus (PMMoV) copies per ml as a ratio (from observed data) by location which can confirm one case with 50% probability.

| Facility | SARS-CoV-2 (N1)/PMMoV | | | Mean of $p_{ts} \geq 0 \cdot 5$ |
|---|---|---|---|---|
| | Minimum | Median | Mean | |
| A | $2{\cdot}2550\times10^{-7}$ | $3{\cdot}0477\times10^{-4}$ | $1{\cdot}4231\times10^{-3}$ | 0·68720 |
| B | $9{\cdot}4500\times10^{-6}$ | $2{\cdot}3350\times10^{-4}$ | $2{\cdot}1445\times10^{-3}$ | 0·70163 |
| C | $1{\cdot}3913\times10^{-6}$ | $6{\cdot}2590\times10^{-4}$ | $1{\cdot}7391\times10^{-3}$ | 0·76319 |
| D | $2{\cdot}0650\times10^{-6}$ | $1{\cdot}6331\times10^{-3}$ | $6{\cdot}2072\times10^{-3}$ | 0·81814 |
| E | $5{\cdot}1500\times10^{-7}$ | $5{\cdot}4854\times10^{-4}$ | $6{\cdot}2694\times10^{-3}$ | 0·77407 |
| F | $2{\cdot}7050\times10^{-6}$ | $1{\cdot}1072\times10^{-3}$ | $3{\cdot}3144\times10^{-2}$ | 0·83129 |
| G | $3{\cdot}5300\times10^{-6}$ | $8{\cdot}6709\times10^{-4}$ | $1{\cdot}6464\times10^{-3}$ | 0·80803 |
| H | $1{\cdot}3085\times10^{-6}$ | $2{\cdot}6386\times10^{-4}$ | $1{\cdot}2609\times10^{-3}$ | 0·77954 |
| I | $8{\cdot}0850\times10^{-7}$ | $6{\cdot}2133\times10^{-4}$ | $2{\cdot}1970\times10^{-3}$ | 0·77555 |
| J | $1{\cdot}0900\times10^{-5}$ | $6{\cdot}8039\times10^{-4}$ | $1{\cdot}2382\times10^{-3}$ | 0·79152 |
| K | $6{\cdot}9125\times10^{-6}$ | $4{\cdot}5700\times10^{-4}$ | $1{\cdot}2733\times10^{-3}$ | 7·22460 |
| L | $4{\cdot}3200\times10^{-7}$ | $3{\cdot}5732\times10^{-5}$ | $3{\cdot}8508\times10^{-4}$ | 0·67637 |
| M | $1{\cdot}0333\times10^{-6}$ | $4{\cdot}0959\times10^{-4}$ | $1{\cdot}9216\times10^{-2}$ | 0·76447 |
| N | $2{\cdot}0067\times10^{-6}$ | $1{\cdot}3448\times10^{-4}$ | $6{\cdot}0338\times10^{-4}$ | 0·75499 |

Table E.5: Selected SARS-CoV-2 (N1) copies per ml normalized by pepper mild mottle virus (PMMoV) copies per ml as a ratio (from observed data) by location which can confirm one case with 60% probability.

| Facility | SARS-CoV-2 (N1)/PMMoV | | | Mean of $p_{ts} \geq 0 \cdot 6$ |
|---|---|---|---|---|
| | Minimum | Median | Mean | |
| A | $9{\cdot}1132\times10^{-5}$ | $1{\cdot}2819\times10^{-3}$ | $2{\cdot}4953\times10^{-3}$ | 0·83125 |
| B | $9{\cdot}4500\times10^{-6}$ | $8{\cdot}0960\times10^{-4}$ | $3{\cdot}1131\times10^{-3}$ | 0·79422 |
| C | $1{\cdot}3913\times10^{-6}$ | $9{\cdot}4546\times10^{-4}$ | $1{\cdot}9298\times10^{-3}$ | 0·79432 |
| D | $2{\cdot}4200\times10^{-5}$ | $2{\cdot}0288\times10^{-3}$ | $6{\cdot}5950\times10^{-3}$ | 0·83799 |
| E | $3{\cdot}1467\times10^{-5}$ | $1{\cdot}3235\times10^{-3}$ | $7{\cdot}6061\times10^{-3}$ | 0·82870 |
| F | $8{\cdot}2350\times10^{-6}$ | $2{\cdot}2208\times10^{-3}$ | $3{\cdot}9712\times10^{-2}$ | 0·88866 |
| G | $2{\cdot}1770\times10^{-5}$ | $1{\cdot}3178\times10^{-3}$ | $2{\cdot}0109\times10^{-3}$ | 0·88219 |
| H | $3{\cdot}4233\times10^{-5}$ | $6{\cdot}3417\times10^{-4}$ | $1{\cdot}6238\times10^{-3}$ | 0·85998 |
| I | $2{\cdot}6100\times10^{-6}$ | $7{\cdot}9227\times10^{-4}$ | $2{\cdot}7765\times10^{-3}$ | 0·84794 |
| J | $1{\cdot}7997\times10^{-5}$ | $1{\cdot}1496\times10^{-3}$ | $1{\cdot}4990\times10^{-3}$ | 0·85966 |
| K | $6{\cdot}9125\times10^{-6}$ | $7{\cdot}3037\times10^{-4}$ | $1{\cdot}8167\times10^{-3}$ | 0·82788 |
| L | $7{\cdot}3000\times10^{-7}$ | $1{\cdot}1959\times10^{-4}$ | $5{\cdot}5338\times10^{-4}$ | 0·74610 |
| M | $1{\cdot}9133\times10^{-5}$ | $1{\cdot}2004\times10^{-3}$ | $2{\cdot}7500\times10^{-2}$ | 0·85983 |
| N | $2{\cdot}0067\times10^{-6}$ | $2{\cdot}4362\times10^{-4}$ | $7{\cdot}1254\times10^{-4}$ | 0·81345 |

Table E.6: Selected SARS-CoV-2 (N1) copies per ml normalized by pepper mild mottle virus (PMMoV) copies per ml as a ratio (from observed data) by location which can confirm one case with 70% probability.

| Facility | SARS-CoV-2 (N1)/PMMoV | | | Mean of $p_{ts} \geq 0 \cdot 7$ |
|---|---|---|---|---|
| | Minimum | Median | Mean | |
| A | $9{\cdot}1132\times10^{-5}$ | $1{\cdot}3276\times10^{-3}$ | $3{\cdot}3686\times10^{-3}$ | 0·89787 |
| B | $2{\cdot}0167\times10^{-5}$ | $1{\cdot}2344\times10^{-3}$ | $4{\cdot}0051\times10^{-3}$ | 0·84870 |
| C | $9{\cdot}5500\times10^{-6}$ | $1{\cdot}4408\times10^{-3}$ | $2{\cdot}6581\times10^{-3}$ | 0·86946 |
| D | $2{\cdot}4200\times10^{-5}$ | $3{\cdot}1539\times10^{-3}$ | $8{\cdot}0506\times10^{-3}$ | 0·88882 |
| E | $3{\cdot}1467\times10^{-5}$ | $1{\cdot}8723\times10^{-3}$ | $1{\cdot}0193\times10^{-2}$ | 0·90719 |
| F | $8{\cdot}2350\times10^{-6}$ | $2{\cdot}8129\times10^{-3}$ | $4{\cdot}1786\times10^{-2}$ | 0·90285 |
| G | $2{\cdot}1770\times10^{-5}$ | $1{\cdot}8279\times10^{-3}$ | $2{\cdot}2887\times10^{-3}$ | 0·91644 |
| H | $3{\cdot}4233\times10^{-5}$ | $1{\cdot}1208\times10^{-3}$ | $1{\cdot}7890\times10^{-3}$ | 0·88785 |
| I | $6{\cdot}6894\times10^{-6}$ | $2{\cdot}0206\times10^{-3}$ | $3{\cdot}4656\times10^{-3}$ | 0·91467 |
| J | $1{\cdot}7997\times10^{-5}$ | $1{\cdot}5287\times10^{-3}$ | $1{\cdot}8216\times10^{-3}$ | 0·93688 |
| K | $8{\cdot}4790\times10^{-5}$ | $1{\cdot}2989\times10^{-3}$ | $2{\cdot}5344\times10^{-3}$ | 0·92419 |
| L | $5{\cdot}1900\times10^{-6}$ | $4{\cdot}8964\times10^{-4}$ | $7{\cdot}5647\times10^{-4}$ | 0·81888 |
| M | $1{\cdot}9133\times10^{-5}$ | $2{\cdot}1806\times10^{-3}$ | $3{\cdot}2924\times10^{-2}$ | 0·90468 |
| N | $2{\cdot}0067\times10^{-6}$ | $3{\cdot}0900\times10^{-4}$ | $8{\cdot}8362\times10^{-4}$ | 0·86775 |

Table E.7: Selected SARS-CoV-2 (N1) copies per ml normalized by pepper mild mottle virus (PMMoV) copies per ml as a ratio (from observed data) by location which can confirm one case with 90% probability.

| Facility | SARS-CoV-2 (N1)/PMMoV | | | Mean of $p_{ts} \geq 0 \cdot 9$ |
|---|---|---|---|---|
| | Minimum | Median | Mean | |
| A | $1{\cdot}3276\times10^{-3}$ | $4{\cdot}6797\times10^{-3}$ | $5{\cdot}1567\times10^{-3}$ | 0·98192 |
| B | $6{\cdot}0593\times10^{-5}$ | $2{\cdot}7099\times10^{-3}$ | $7{\cdot}9721\times10^{-3}$ | 0·96334 |
| C | $4{\cdot}4191\times10^{-4}$ | $4{\cdot}1305\times10^{-3}$ | $4{\cdot}6130\times10^{-3}$ | 0·96026 |
| D | $2{\cdot}4246\times10^{-3}$ | $7{\cdot}4374\times10^{-3}$ | $1{\cdot}2727\times10^{-2}$ | 0·98632 |
| E | $1{\cdot}6570\times10^{-4}$ | $3{\cdot}8532\times10^{-3}$ | $1{\cdot}5032\times10^{-2}$ | 0·96987 |
| F | $8{\cdot}6723\times10^{-4}$ | $7{\cdot}1838\times10^{-3}$ | $6{\cdot}5746\times10^{-2}$ | 0·97470 |
| G | $3{\cdot}4976\times10^{-4}$ | $2{\cdot}6793\times10^{-3}$ | $2{\cdot}8130\times10^{-3}$ | 0·96956 |
| H | $8{\cdot}3551\times10^{-5}$ | $2{\cdot}3507\times10^{-3}$ | $2{\cdot}7863\times10^{-3}$ | 0·95608 |
| I | $9{\cdot}9748\times10^{-5}$ | $3{\cdot}0372\times10^{-3}$ | $4{\cdot}8338\times10^{-3}$ | 0·96822 |
| J | $4{\cdot}7200\times10^{-5}$ | $2{\cdot}0477\times10^{-3}$ | $2{\cdot}2551\times10^{-3}$ | 0·97912 |
| K | $1{\cdot}3570\times10^{-4}$ | $1{\cdot}4225\times10^{-3}$ | $3{\cdot}0316\times10^{-3}$ | 0·95641 |
| L | $3{\cdot}4938\times10^{-4}$ | $7{\cdot}3263\times10^{-4}$ | $1{\cdot}3915\times10^{-3}$ | 0·94086 |
| M | $9{\cdot}0293\times10^{-5}$ | $4{\cdot}3314\times10^{-3}$ | $4{\cdot}8042\times10^{-2}$ | 0·96245 |
| N | $1{\cdot}1576\times10^{-4}$ | $7{\cdot}7051\times10^{-4}$ | $1{\cdot}3805\times10^{-3}$ | 0·95013 |

Table E.8: Selected SARS-CoV-2 (N1) copies per ml normalized by pepper mild mottle virus (PMMoV) copies per ml as a ratio (from observed data) by location which can confirm one case with 95% probability.

| Facility | SARS-CoV-2 (N1)/PMMoV | | | Mean of $p_{ts} \geq 0 \cdot 95$ |
|---|---|---|---|---|
| | Minimum | Median | Mean | |
| A | $1{\cdot}3276{\times}10^{-3}$ | $4{\cdot}6797{\times}10^{-3}$ | $5{\cdot}1567{\times}10^{-3}$ | 0·98192 |
| B | $1{\cdot}2344{\times}10^{-3}$ | $6{\cdot}3462{\times}10^{-3}$ | $1{\cdot}0755{\times}10^{-2}$ | 0·97889 |
| C | $1{\cdot}4408{\times}10^{-3}$ | $4{\cdot}6549{\times}10^{-3}$ | $5{\cdot}5934{\times}10^{-3}$ | 0·98170 |
| D | $2{\cdot}4246{\times}10^{-3}$ | $7{\cdot}4374{\times}10^{-3}$ | $1{\cdot}2727{\times}10^{-2}$ | 0·98632 |
| E | $3{\cdot}3796{\times}10^{-4}$ | $4{\cdot}5999{\times}10^{-3}$ | $1{\cdot}9319{\times}10^{-2}$ | 0·98202 |
| F | $1{\cdot}6287{\times}10^{-3}$ | $8{\cdot}9371{\times}10^{-3}$ | $8{\cdot}7049{\times}10^{-2}$ | 0·99228 |
| G | $3{\cdot}4976{\times}10^{-4}$ | $2{\cdot}8302{\times}10^{-3}$ | $2{\cdot}8328{\times}10^{-3}$ | 0·97475 |
| H | $8{\cdot}3551{\times}10^{-5}$ | $3{\cdot}0363{\times}10^{-3}$ | $3{\cdot}5807{\times}10^{-3}$ | 0·97362 |
| I | $1{\cdot}1784{\times}10^{-3}$ | $3{\cdot}2910{\times}10^{-3}$ | $6{\cdot}2833{\times}10^{-3}$ | 0·98828 |
| J | $4{\cdot}7200{\times}10^{-5}$ | $2{\cdot}0477{\times}10^{-3}$ | $2{\cdot}2551{\times}10^{-3}$ | 0·97912 |
| K | $1{\cdot}3570{\times}10^{-4}$ | $1{\cdot}4225{\times}10^{-3}$ | $3{\cdot}4036{\times}10^{-3}$ | 0·97406 |
| L | $7{\cdot}2997{\times}10^{-4}$ | $1{\cdot}3379{\times}10^{-3}$ | $1{\cdot}8779{\times}10^{-3}$ | 0·96496 |
| M | $8{\cdot}9398{\times}10^{-4}$ | $4{\cdot}8779{\times}10^{-3}$ | $6{\cdot}7554{\times}10^{-2}$ | 0·98123 |
| N | $6{\cdot}0025{\times}10^{-4}$ | $1{\cdot}2014{\times}10^{-3}$ | $1{\cdot}9938{\times}10^{-3}$ | 0·97232 |

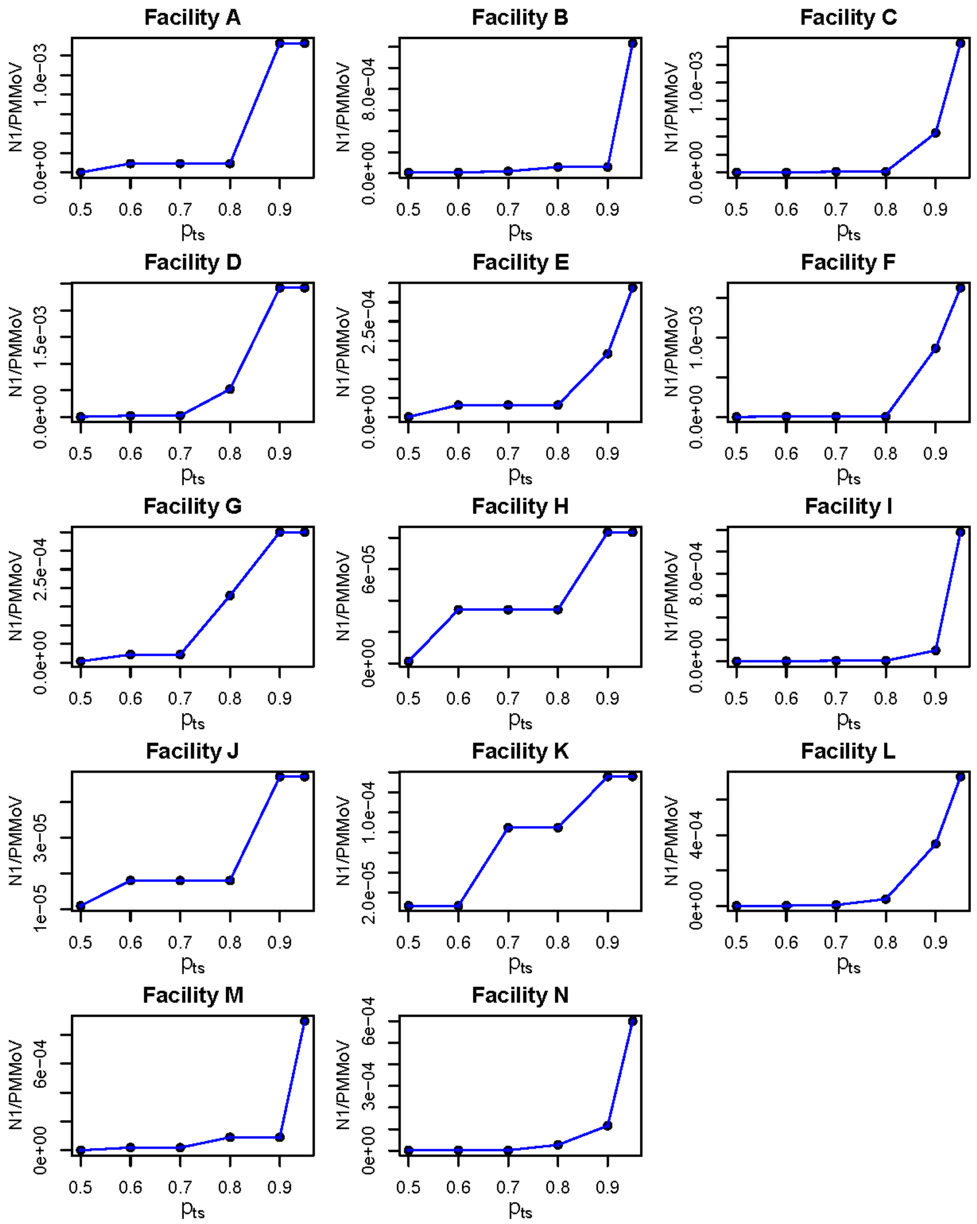

Figure E.4: The minimum value of SARS-CoV-2 (N1) copies per ml normalized by pepper mild mottle virus (PMMoV) copies per ml as a ratio associated with at least one case for different thresholds of reporting probabilities ($p_{ts}$) at each facility.

### Appendix F. Prediction Result

Table F.9: Summary of prediction results for Facility G, H, I during Feb 27th-March 12th, 2022.

| Facility | Week (week number) | Observed $Z_{ts}$ | SARS-CoV-2 (N1) / PMMoV | 95% C.I of $p_{ts}$ | | Predicted mean of $p_{ts}$ |
|---|---|---|---|---|---|---|
| | | | | $2 \cdot 5\%$ | $97 \cdot 5\%$ | |
| G | 02/27/2022 - 03/05/2022 (57) | 22 | $6 \cdot 31 \times 10^{-4}$ | 0·863 | 0·903 | 0·884 |
| | 03/06/2022 - 03/12/2022 (58) | 0 | $2 \cdot 19 \times 10^{-5}$ | 0·600 | 0·696 | 0·649 |
| H | 02/27/2022 - 03/05/2022 (57) | 2 | $3 \cdot 57 \times 10^{-5}$ | 0·714 | 0·787 | 0·752 |
| | 03/06/2022 - 03/12/2022 (58) | 0 | $2 \cdot 45 \times 10^{-5}$ | 0·546 | 0·645 | 0·596 |
| I | 02/27/2022 - 03/05/2022 (57) | 1 | $6 \cdot 47 \times 10^{-6}$ | 0·523 | 0·618 | 0·571 |
| | 03/06/2022 - 03/12/2022 (58) | 1 | $2 \cdot 61 \times 10^{-6}$ | 0·461 | 0·560 | 0·511 |

Table F.10: Summary of prediction results for Facility C, H, L during Feb 5th-15th, 2023.

| Facility | Week (week number) | Observed $Z_{ts}$ | SARS-CoV-2 (N1) / PMMoV | 95% C.I of $p_{ts}$ | | Predicted mean of $p_{ts}$ |
|---|---|---|---|---|---|---|
| | | | | $2 \cdot 5\%$ | $97 \cdot 5\%$ | |
| C | 02/05/2023 - 02/11/2023 (106) | 5 | $1 \cdot 13 \times 10^{-3}$ | 0·711 | 0·784 | 0·75 |
| | 02/12/2023 - 02/18/2023 (107) | 14 | $4 \cdot 65 \times 10^{-3}$ | 0·989 | 0·992 | 0·99 |
| | 02/19/2023 - 02/15/2023 (108) | 3 | $9 \cdot 05 \times 10^{-4}$ | 0·670 | 0·750 | 0·71 |
| H | 02/05/2023 - 02/11/2023 (106) | 9 | $2 \cdot 35 \times 10^{-3}$ | 0·907 | 0·935 | 0·92 |
| | 02/12/2023 - 02/18/2023 (107) | 1 | $1 \cdot 40 \times 10^{-3}$ | 0·775 | 0·838 | 0·81 |
| | 02/19/2023 - 02/15/2023 (108) | 0 | $1 \cdot 36 \times 10^{-4}$ | 0·485 | 0·587 | 0·54 |
| L | 02/05/2023 - 02/11/2023 (106) | 0 | $3 \cdot 67 \times 10^{-3}$ | 0·979 | 0·986 | 0·98 |
| | 02/12/2023 - 02/18/2023 (107) | 0 | $1 \cdot 32 \times 10^{-3}$ | 0·776 | 0·841 | 0·81 |
| | 02/19/2023 - 02/15/2023 (108) | 30 | $4 \cdot 38 \times 10^{-4}$ | 0·569 | 0·668 | 0·62 |

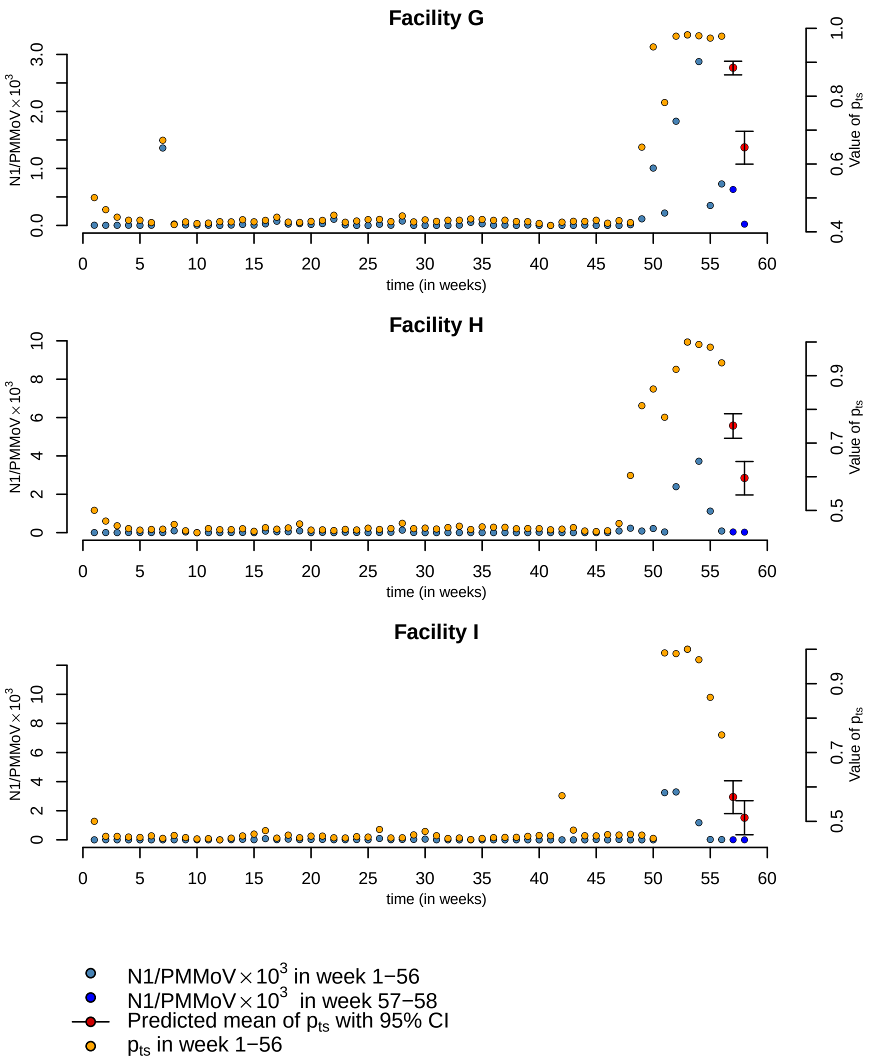

Figure F.5: The figure illustrates the predicted probabilities $p_{ts}$ for weeks 57-58 (highlighted in red circles) by utilizing the SARS-CoV-2 (N1) copies per ml normalized by pepper mild mottle virus (PMMoV) copies per ml as a ratio from weeks 57-58 (green circles) without prior knowledge of the reported positive cases, along with the mean estimated $p_{ts}$ from the training dataset for weeks 1-56 (yellow circles).

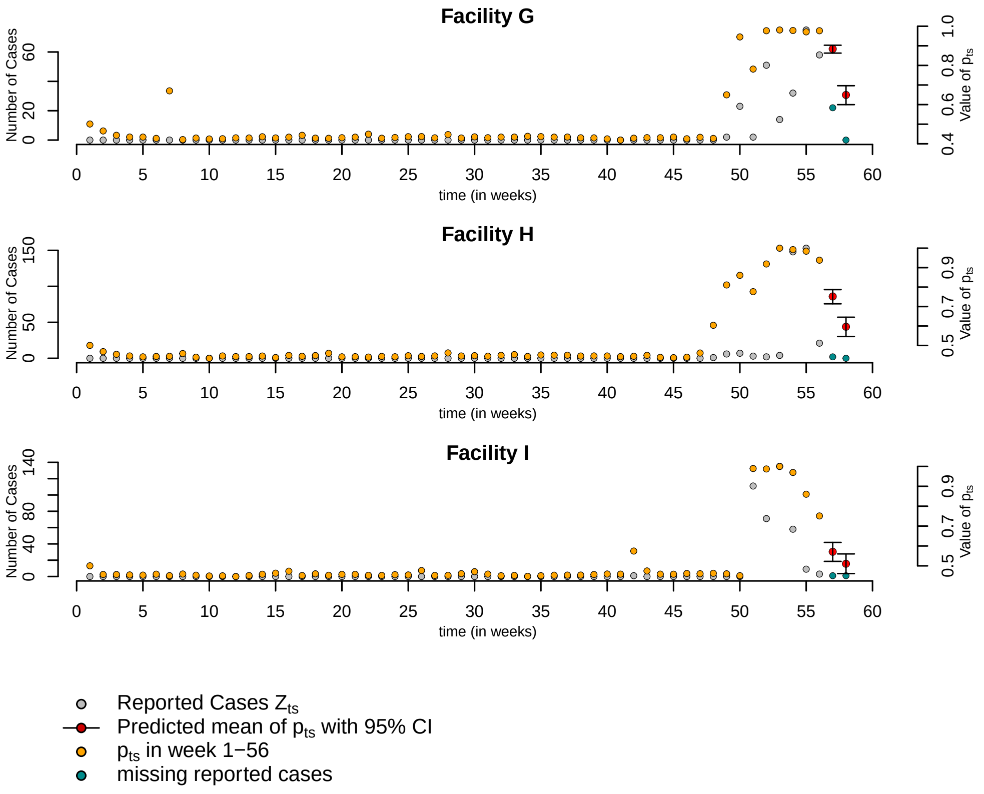

Figure F.6: The figure illustrates the predicted probabilities $p_{ts}$ for weeks 57-58 (highlighted in red circles) by utilizing the SARS-CoV-2 (N1) copies per ml normalized by pepper mild mottle virus (PMMoV) copies per ml as a ratio from weeks 57-58 (green circles) without prior knowledge of the reported positive cases, along with the reported $Z_{ts}$ from the training dataset in weeks 1-56 (grey circles) and in prediction dataset in week 57-58 (green circles).

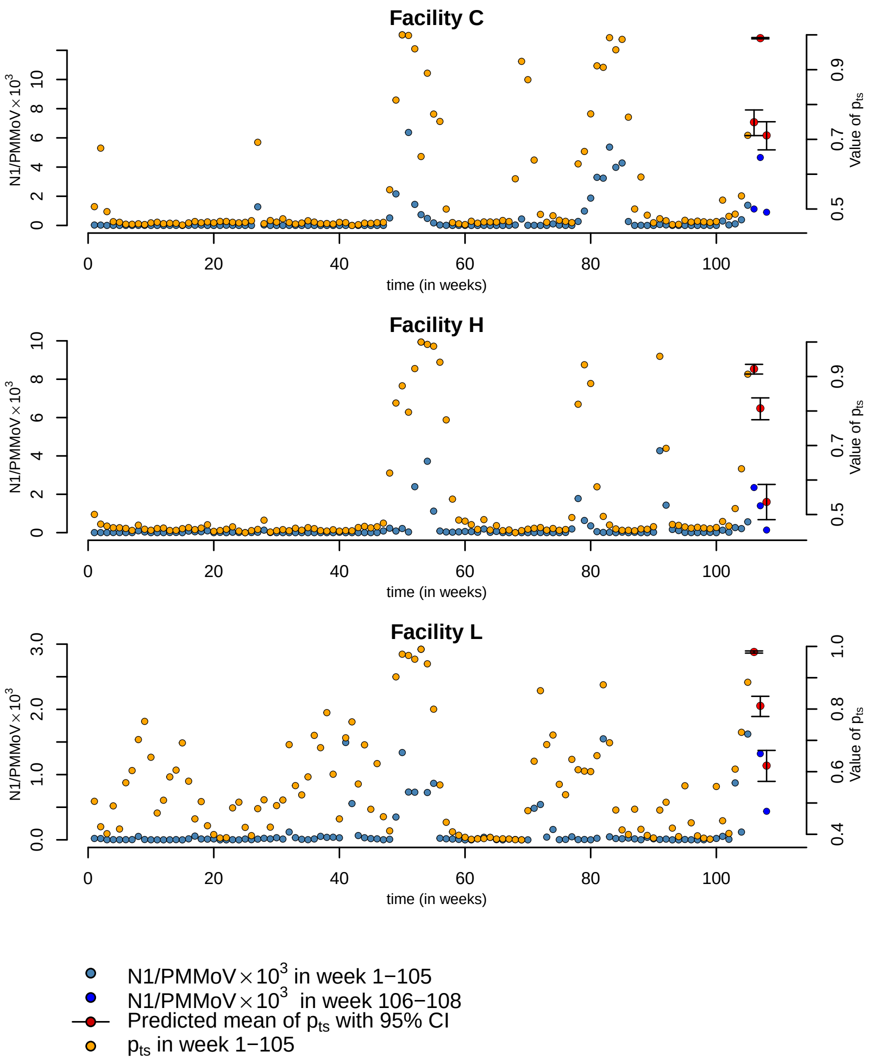

Figure F.7: The figure illustrates the predicted probabilities $p_{ts}$ for weeks 106-108 (highlighted in red circles) by utilizing the SARS-CoV-2 (N1) copies per ml normalized by pepper mild mottle virus (PMMoV) copies per ml as a ratio from week 106-108 (green circles) without prior knowledge of the reported positive cases, along with the mean estimated $p_{ts}$ from the training dataset in weeks 1-105 (yellow circles).

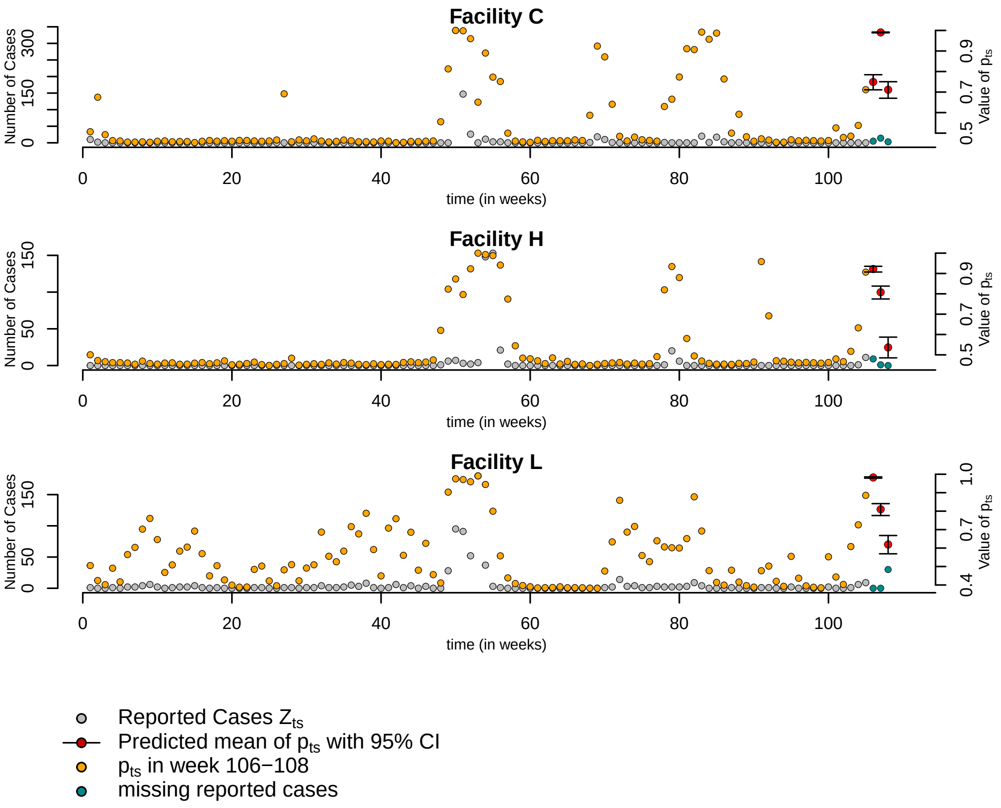

Figure F.8: The figure illustrates the predicted probabilities $p_{ts}$ for weeks 106-108 (highlighted in red circles) by utilizing the SARS-CoV-2 (N1) copies per ml normalized by pepper mild mottle virus (PMMoV) copies per ml as a ratio from weeks 106-108 (green circles) without prior knowledge of the reported positive cases, along with the reported $Z_{ts}$ from the training dataset in weeks 1-105 (grey circles) and in prediction dataset in week 106-108 (green circles).

### Appendix G. Sensitivity Analysis

All model parameters have specified prior distributions. This analysis focuses on the two most critical model parameters: the variance of the prior for $a_s$ that impacts the true arrival intensity of true positive cases: $\sigma^2_{a_{0,s}}$, and the covariance matrix of the prior for $\vec{\beta}_s$ that impacts the reporting probability: $P_{\beta_0}$. In this sensitivity analysis, we explore different prior specifications to investigate how various priors, which may influence the final thresholds of

SARS-CoV-2 (N1)/PMMoV ratio.

Typically, prior informativeness falls into three categories: informative, weakly informative, and diffuse. Informative priors contain a substantial amount of information about a specific parameter, leading to a high probability mass concentrated within a relatively narrow range of possible values. Weakly informative priors allow for more variation or spread compared to informative priors. On the other hand, diffuse priors offer minimal to no information about the parameter value.

For this study, we assign informative priors $N(0, 0\cdot 1)$, weakly informative priors $N(0, 1)$, and diffuse priors $N(0, 1000)$ to $a_s$ and $\vec{\beta}_s$, respectively. This results in nine different conditions of priors that we analyze to understand their impact on the thresholds of SARS-CoV-2 (N1)/PMMoV ratio:

(1) $\sigma^2_{a_{0,s}} = 0 \cdot 1 \quad , \quad P_{\beta_0} = 0 \cdot 1\, \mathbb{I}_2$
(2) $\sigma^2_{a_{0,s}} = 0 \cdot 1 \quad , \quad P_{\beta_0} = \mathbb{I}_2$
(3) $\sigma^2_{a_{0,s}} = 0 \cdot 1 \quad , \quad P_{\beta_0} = 1000\, \mathbb{I}_2$
(4) $\sigma^2_{a_{0,s}} = 1 \quad , \quad P_{\beta_0} = 0 \cdot 1\, \mathbb{I}_2$
(5) $\sigma^2_{a_{0,s}} = 1 \quad , \quad P_{\beta_0} = \mathbb{I}_2$
(6) $\sigma^2_{a_{0,s}} = 1 \quad , \quad P_{\beta_0} = 1000\, \mathbb{I}_2$
(7) $\sigma^2_{a_{0,s}} = 1000 \quad , \quad P_{\beta_0} = 0 \cdot 1\, \mathbb{I}_2$
(8) $\sigma^2_{a_{0,s}} = 1000 \quad , \quad P_{\beta_0} = \mathbb{I}_2$
(9) $\sigma^2_{a_{0,s}} = 1000 \quad , \quad P_{\beta_0} = 1000\, \mathbb{I}_2$

where $\sigma^2_{a_{0,s}}$ is the variance of $a_s$, $P_{\beta_0}$ is the variance of the vector $\vec{\beta}_s$, and $\mathbb{I}_2$ is the $2 \times 2$ identity matrix.

We determine the SARS-CoV-2 (N1)/PMMoV ratio corresponding to $p_{ts} \geq 0 \cdot 8$ and compute the associated summary statistics, including the minimum, median, and mean. The summarized results are presented in the following table, with the respective condition numbers written in the parentheses:

Table G.11: Sensitivity Analysis Table

<table>
<tr><th rowspan="2">Facility</th><th colspan="3">SARS-CoV-2 (N1)/PMMoV (combination)</th></tr>
<tr><th>Minimum</th><th>Median</th><th>Mean</th></tr>
<tr><td rowspan="2">A</td><td>$9 \cdot 1100 \times 10^{-5}$ (1,2,3,5,6,8,9)</td><td>$3 \cdot 0037 \times 10^{-3}$ (1,2,3,5,6,8,9)</td><td>$3 \cdot 8903 \times 10^{-3}$ (1,2,3,5,6,8,9)</td></tr>
<tr><td>$1 \cdot 3276 \times 10^{-3}$ (4,7)</td><td>$4 \cdot 6797 \times 10^{-3}$ (4,7)</td><td>$5 \cdot 1567 \times 10^{-3}$ (4,7)</td></tr>
<tr><td rowspan="3">B</td><td rowspan="3">$6 \cdot 0600 \times 10^{-5}$ (1 - 9)</td><td>$2 \cdot 3396 \times 10^{-3}$ (1,2,4,7)</td><td>$7 \cdot 0251 \times 10^{-3}$ (1,2,4,7)</td></tr>
<tr><td rowspan="2">$1 \cdot 9692 \times 10^{-3}$ (3,5,6,8,9)</td><td>$6 \cdot 3162 \times 10^{-3}$ (3,6,9)</td></tr>
<tr><td>$6 \cdot 3818 \times 10^{-3}$ (5,8)</td></tr>
<tr><td rowspan="2">C</td><td rowspan="2">$9 \cdot 5500 \times 10^{-6}$ (1 - 9)</td><td>$3 \cdot 2386 \times 10^{-3}$ (1,4,7)</td><td>$3 \cdot 3877 \times 10^{-3}$ (1,4,7)</td></tr>
<tr><td>$2 \cdot 7007 \times 10^{-3}$ (2,3,5,6,8,9)</td><td>$3 \cdot 2930 \times 10^{-3}$ (2,3,5,6,8,9)</td></tr>
<tr><td rowspan="2">D</td><td>$2 \cdot 4246 \times 10^{-3}$ (1,4,5,7)</td><td>$7 \cdot 4374 \times 10^{-3}$ (1,4,5,7)</td><td>$1 \cdot 2727 \times 10^{-2}$ (1,4,5,7)</td></tr>
<tr><td>$5 \cdot 2148 \times 10^{-4}$ (2,3,6,8,9)</td><td>$6 \cdot 5108 \times 10^{-3}$ (2,3,6,8,9)</td><td>$1 \cdot 1371 \times 10^{-2}$ (2,3,6,8,9)</td></tr>
<tr><td rowspan="3">E</td><td rowspan="3">$3 \cdot 1500 \times 10^{-5}$ (1 - 9)</td><td>$2 \cdot 2117 \times 10^{-3}$ (1,7)</td><td>$1 \cdot 1788 \times 10^{-2}$ (1,7)</td></tr>
<tr><td>$1 \cdot 9448 \times 10^{-3}$ (2,3,5,6,9)</td><td>$1 \cdot 0657 \times 10^{-2}$ (2,3,5,6,9)</td></tr>
<tr><td>$2 \cdot 0783 \times 10^{-3}$ (4,8)</td><td>$1 \cdot 1241 \times 10^{-2}$ (4,8)</td></tr>
<tr><td rowspan="2">F</td><td rowspan="2">$8 \cdot 2400 \times 10^{-6}$ (1 - 9)</td><td>$4 \cdot 2831 \times 10^{-3}$ (1,2,4,5,7,8)</td><td>$4 \cdot 9519 \times 10^{-2}$ (1,2,4,5,7,8)</td></tr>
<tr><td>$3 \cdot 2922 \times 10^{-3}$ (3,6,9)</td><td>$4 \cdot 6656 \times 10^{-2}$ (3,6,9)</td></tr>
<tr><td>G</td><td>$1 \cdot 7996 \times 10^{-4}$ (1 - 9)</td><td>$2 \cdot 5285 \times 10^{-3}$ (1 - 9)</td><td>$2 \cdot 6581 \times 10^{-3}$ (1 - 9)</td></tr>
</table>

Table G.11: Sensitivity Analysis Table

| Facility | SARS-CoV-2 (N1)/PMMoV (combination) | | |
|---|---|---|---|
| | Minimum | Median | Mean |
| H | $8 \cdot 3600 \times 10^{-5}$ (1,4,7) | $1 \cdot 2616 \times 10^{-3}$ (1,4,7) | $2 \cdot 0650 \times 10^{-3}$ (1,4,7) |
| | $3 \cdot 4200 \times 10^{-5}$ (2,3,5,6,8,9) | $1 \cdot 1208 \times 10^{-3}$ (2,6,9) | $1 \cdot 9296 \times 10^{-3}$ (2,6,9) |
| | | $8 \cdot 7746 \times 10^{-4}$ (3,5,8) | $1 \cdot 8113 \times 10^{-3}$ (3,5,8) |
| I | $2 \cdot 2800 \times 10^{-5}$ (1,4,7) | $2 \cdot 2221 \times 10^{-3}$ (1,4,7) | $4 \cdot 0314 \times 10^{-3}$ (1,4,7) |
| | $6 \cdot 6900 \times 10^{-6}$ (2,3,5,6,8,9) | $2 \cdot 1556 \times 10^{-3}$ (2,3,5,6,8,9) | $3 \cdot 6684 \times 10^{-3}$ (2,3,5,6,8,9) |
| J | $1 \cdot 800 \times 10^{-5}$ (1 - 9) | $1 \cdot 6031 \times 10^{-3}$ (1 - 9) | $1 \cdot 8550 \times 10^{-3}$ (1 - 9) |
| K | $8 \cdot 480 \times 10^{-5}$ (1 - 9) | $1 \cdot 2989 \times 10^{-3}$ (1 - 9) | $2 \cdot 5344 \times 10^{-3}$ (1 - 9) |
| L | $3 \cdot 8200 \times 10^{-5}$ (1,2,4,5,8) | $7 \cdot 3130 \times 10^{-4}$ (1,4) | $1 \cdot 1481 \times 10^{-3}$ (1,4) |
| | | $7 \cdot 3263 \times 10^{-4}$ (2,5,8) | $1 \cdot 1936 \times 10^{-3}$ (2,5,8) |
| | $3 \cdot 4938 \times 10^{-4}$ (3,6,9) | $7 \cdot 3263 \times 10^{-4}$ (3,6) | $1 \cdot 3285 \times 10^{-3}$ (3,6) |
| | | $7 \cdot 9929 \times 10^{-4}$ (9) | $1 \cdot 2899 \times 10^{-3}$ (9) |
| | $5 \cdot 1900 \times 10^{-6}$ (7) | $7 \cdot 2997 \times 10^{-4}$ (7) | $1 \cdot 0719 \times 10^{-3}$ (7) |
| M | $9 \cdot 0300 \times 10^{-5}$ (1 - 9) | $3 \cdot 4089 \times 10^{-3}$ (1,2,3,4,6,7) | $3 \cdot 9120 \times 10^{-2}$ (1,2,3,4,6,7) |
| | | $2 \cdot 8076 \times 10^{-3}$ (5,8) | $3 \cdot 7362 \times 10^{-2}$ (5,8) |
| | | $2 \cdot 2064 \times 10^{-3}$ (9) | $3 \cdot 5772 \times 10^{-2}$ (9) |
| N | $2 \cdot 6800 \times 10^{-5}$ (1 - 9) | $5 \cdot 6938 \times 10^{-4}$ (1 - 9) | $1 \cdot 1400 \times 10^{-3}$ (1 - 9) |

Based on the findings presented in Table (G.11), a sensitivity analysis was conducted to assess the impact of different prior conditions on the results. Out of the 14 facilities examined, the majority (9 out of 14) consistently showed similar outcomes concerning the minimum SARS-CoV-2 (N1)/PMMoV ratio corresponding to at least one positive case with a probability greater than 80%. This suggests a degree of robustness in the results across these facilities.

In addition to the consistent findings regarding the minimum SARS-CoV-2 (N1)/PMMoV ratio corresponding to at least one positive case with a probability greater than 80% for the majority of facilities, the sensitivity analysis also revealed potential variations in the median of SARS-CoV-2 (N1)/PMMoV ratio under different prior conditions, especially in conditions 1, 4, and 7, where the covariance matrix $P_{\beta_0}$ is highly informative. This variability in the median statistic may be attributed to the significant fluctuations in heterogeneous SARS-CoV-2 (N1)/PMMoV ratios observed in these conditions. This observation highlights the sensitivity of the median statistic to the choice of prior. It was noted that imposing hard constraints may limit the ability to express sufficient uncertainty about the available information regarding the reporting probability. Consequently, it is crucial to interpret the results in light of these constraints and their potential influence on the overall conclusions.

However, despite the variations in median values, a notable finding is that the mean of SARS-CoV-2 (N1)/PMMoV ratio corresponding to at least one positive case with a probability greater than 80% remains consistent and close to each other across these nine different prior conditions. This suggests that the mean can serve as a more stable and reliable measure to focus on when comparing the outcomes under various prior conditions.

In conclusion, the sensitivity analysis highlights the importance of considering different prior conditions and their impact on the results. It emphasizes the need for cautious interpretation, especially when dealing with highly informative priors and hard constraints, and underscores the usefulness of the mean as a robust measure for assessing SARS-CoV-2 (N1)/PMMoV ratios corresponding to positive cases with a probability greater than 80% under these different prior scenarios.

The choice of an alternative prior specification can impact the convergence of parameters in the model. Therefore, it is essential to assess model convergence, even if the original prior specification showed no convergence issues. In the present study, the convergence speed and required iteration number vary across locations based on the current data and experiment.

To ensure convergence, users typically determine the length of the burn-in period using statistical diagnostics while considering the model complexity. If convergence is not achieved for a specific model parameter, practitioners can try doubling or increasing the number of iterations to see if a longer chain resolves the issue.

However, if non-convergence persists, it may indicate that the selected prior is not well-suited for the model or likelihood. In the context of a sensitivity analysis, such results could suggest evidence against choosing that particular prior, given the current model and likelihood.

In the case of these nine different prior conditions in our current experiment, we find that for all locations, the covariance matrix for $\vec{\beta_s}$ needs to be either weakly informative or informative to achieve faster convergence.

### Appendix H. Algorithm

The complete algorithm for the Hierarchical Spatial-temporal Model with Lags is summarized in Algorithm 1. The prediction algorithm is summarized in Algorithm 2.

**Algorithm 1** Hierarchical Spatial-temporal Model with Lags

**Required:**(1) Set value of $\sigma_{\varepsilon_{0,s}}$, $\sigma_{e_{0,s}}$, $\sigma_{a_{0,1}}$, $P_{\beta_0}$, $\mu_{a_{0,s}}$, and $\mu_{\varepsilon_{0,s}}$, as well as provide value of $h$, the shape parameter of $\lambda_{ts}$; (2) Matrix of recorded cases $Z_{ts}$ for all locations $s$; (3) $X_{ts}^{(1)}$ and $X_{ts}^{(2)}$, the design matrix containing factors which influence the reporting probabilities and true positive cases, respectively.

**for** each location $s \in \{1, \cdots, J\}$ **do**

1. Generate $\vec{\varepsilon_s} \sim Normal\left(\mu_{\varepsilon_{0,s}}, \sigma_{\varepsilon_{0,s}}\right)$ and $e_{ts} \sim Normal\left(0, \sigma_{e_{0,s}}\right)$
2. Set $\gamma_{ts} = a_s\, \gamma_{t-1,s} + e_{ts}$
3. Set $\psi_s = X_s^{(1)} \vec{\beta_s} + \vec{\varepsilon_s}$ and generate $\omega_s^{(1)} \sim \mathcal{PG}\left(N_{ts}, \psi_{ts}\right)$
4. Compute $V_{\beta_s}$ and $\mu_{\beta_s}$ following equations (C.25) and (C.26) and estimate $\vec{\beta_s}$ from $Normal\left(\mu_{\beta_s}, V_{\beta_s}\right)$
5. Compute $V_{\varepsilon_s}$ and $\mu_{\varepsilon_s}$ following equations (C.30, C.31) and update $\vec{\varepsilon_s}$ from $Normal\left(\mu_{\varepsilon_s}, V_{\varepsilon_s}\right)$
6. Calculate the reporting probabilities $\pi_{ts}$
7. Set $\phi_{ts} = X_{ts}^{(2)} + \gamma_{ts}$ and draw $\omega_s^{(2)} \sim \mathcal{PG}\left(N_{ts} + h, \phi_{ts}\right)$
8. Compute $V_{a_s}$ and $\mu_{a_s}$ following equations (C.11) and (C.12) and estimate $a_s$ from $Normal\left(\mu_{a_s}, V_{a_s}\right)$
9. Compute $V_{e_s}$ and $\mu_{e_s}$ following equations (C.16, C.17) and update $e_s$ from $Normal\left(\mu_{e_s}, V_{e_s}\right)$
10. Update $\gamma_{ts}$ and calculate $p_{ts}$, the probabilities associated with $N_{ts}$
11. Sample $\lambda_{ts} \sim \mathcal{G}amma\left(N_{ts} + h, p_{ts}\right)$ and $N_{ts} \sim Poisson\left(\left(1 - \pi_{ts}\right)\lambda_{ts}\right)$

**end for**

**Algorithm 2** Prediction model

**Required:** (1) The training results obtained from the Hierarchical Spatio-temporal model with lags; (2) New $X_{ts}^{(1)}$, the design matrix containing factors which influence the reporting probabilities; and (3) New $X_{ts}^{(2)}$, the design matrix containing factors which influence the true positive cases.

**for** each location $s \in \{1, \cdots, J\}$ **do**

1. Compute the estimated coefficients for $a_s$ and $\vec{\beta_s}$ using the results from Algorithm (1).
2. Generate $\vec{\varepsilon_s} \sim Normal\,(0, 0 \cdot 1)$ and $e_{ts} \sim Normal\,(0, 0 \cdot 1)$
3. Calculate reporting probabilities $\pi_{ts} = \frac{e^{\psi_s}}{1+e^{\psi_s}}$, where $\psi_s = X_s^{(1)}\vec{\beta_s} + \vec{\varepsilon_s}$
4. Compute $\gamma_{1s}$ and $\gamma_{ts}$ using the fixed coefficients $a_s$ and $\vec{\beta_s}$ as $\gamma_{1s} = a_s\,\gamma_{-1,s} + e_{ts}$ and $\gamma_{ts} = a_s\,\gamma_{t-1,s} + e_{ts}$, where $\gamma_{-1,s}$ is the estimated value of the last time point in the training model
5. Set $\phi_{ts} = X_{ts}^{(2)} + \gamma_{ts}$ and calculate the probabilities $p_{ts} = \frac{e^{\phi_{ts}}}{1+e^{\phi_{ts}}}$
6. Sample $\lambda_{ts} \sim \mathcal{G}amma\left(h, \frac{p_{ts}}{1-p_{ts}}\right)$ and set $\lambda_{1,s} = \max\{\lambda_{-1,s}, \lambda_{1,s}\}$, where $\lambda_{-1,s}$ is the last time point in the training data set. This is done to accurately capture the number of true cases
7. Generate $Z_{ts} \sim Poisson\,(\pi_{ts}\lambda_{ts})$ and sample $N_{ts} \sim NB\left(h, \frac{1-p_{ts}}{1-\pi_{ts}p_{ts}}\right)$

**end for**

### Appendix I. Model Convergence

A crucial aspect in evaluating the efficacy of our model lies in examining the convergence of its coefficients. The convergence behavior of the coefficients $\beta_1$, $\beta_2$ and $a_s$ is demonstrated in the Appendix, Figure (I.9) and Figure (I.10), which depicts the final 10,000 iterations from a total of 40,000 iterations conducted using the Markov Chain Monte Carlo (MCMC) algorithm. These convergence plots provide evidence that the MCMC algorithm has successfully converged sufficiently fast, indicating stable and reliable estimates for the model coefficients.

Figure I.9: Last 10000 iterations of $\vec{\beta_s}$ by location showing the convergence of the estimated coefficients.

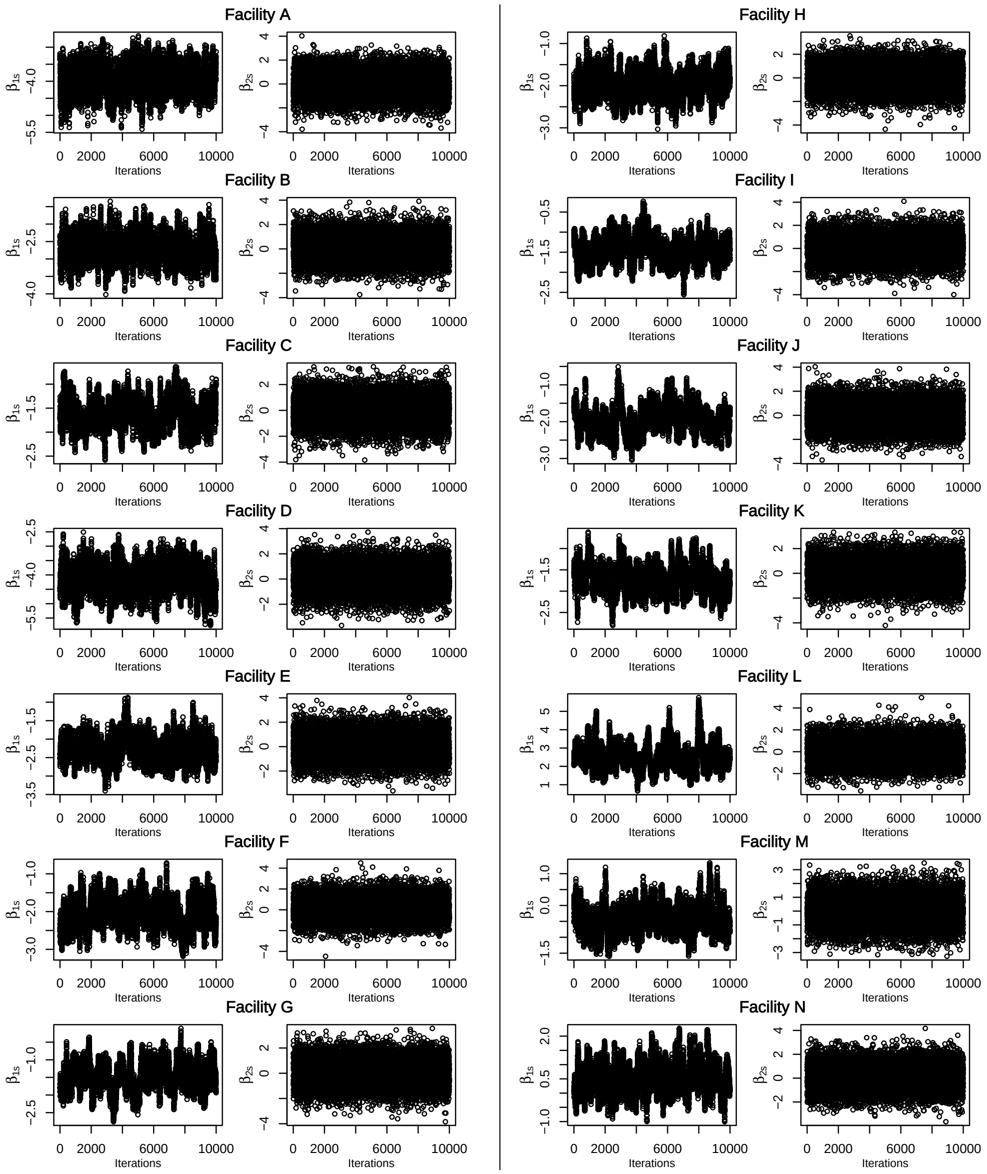

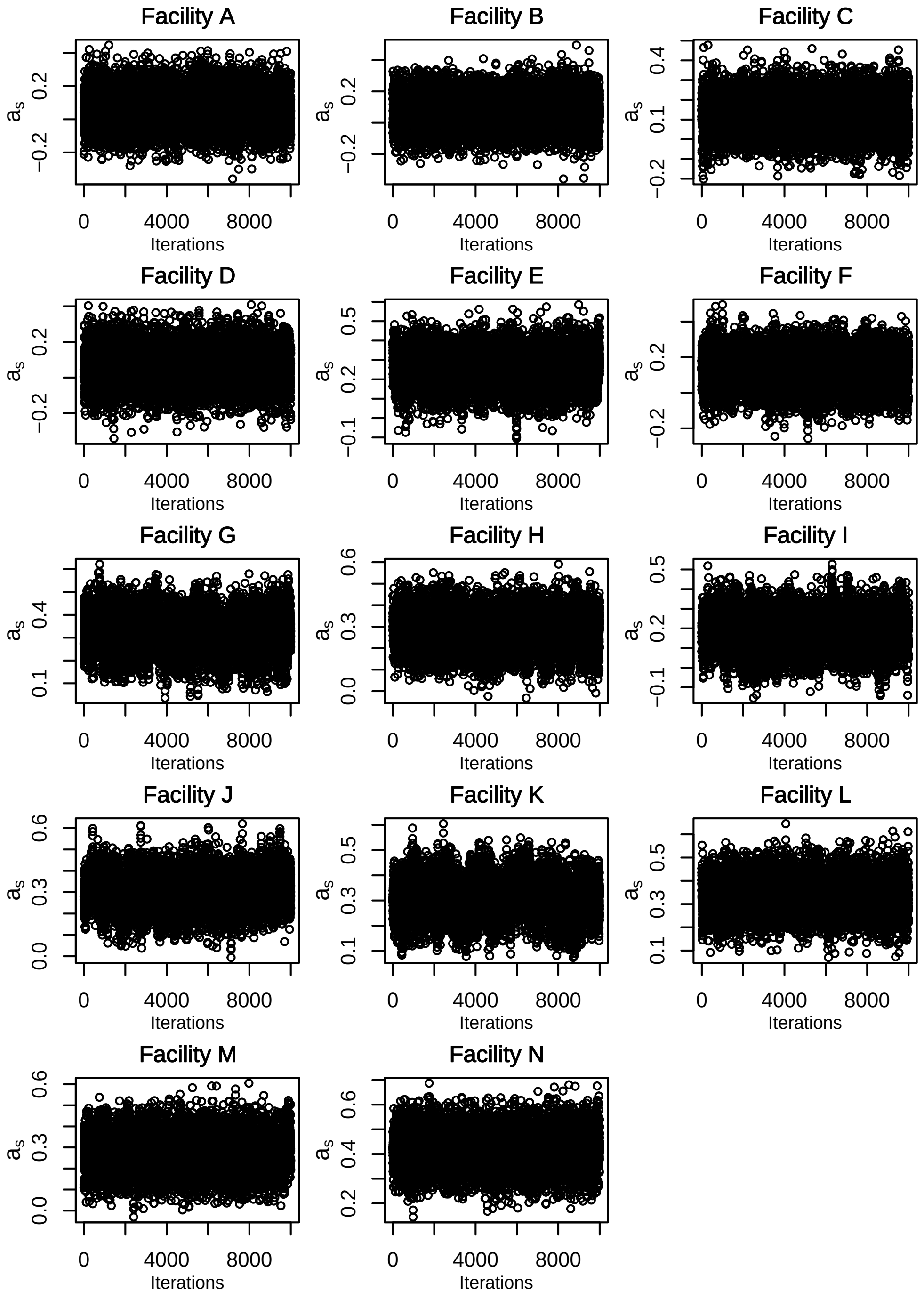

Figure I.10: Last 10000 iterations of $a_s$ by location showing the convergence of the estimated coefficient.

### Appendix J. SARS-CoV-2 (N1) Model

The following figures and tables are the results obtained with the unnormalized SARS-CoV-2 (N1) virus concentration.

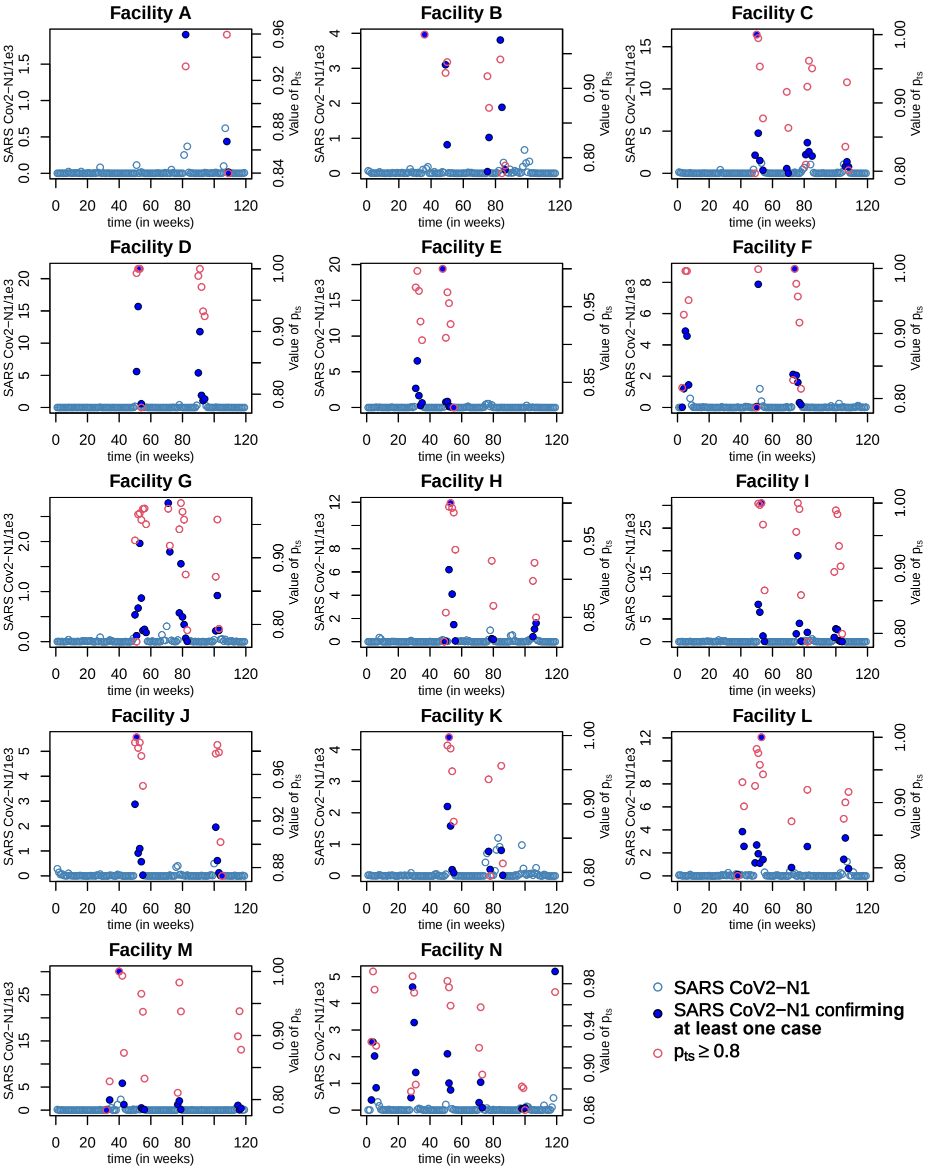

Figure J.11: Value of SARS-CoV-2 (N1) throughout time for all locations, with SARS-CoV-2 (N1) concentration that confirm at least one positive case with 80% probability circled in red. The corresponding $p_{ts} \geq 0 \cdot 8$ is also plotted in red for comparison.

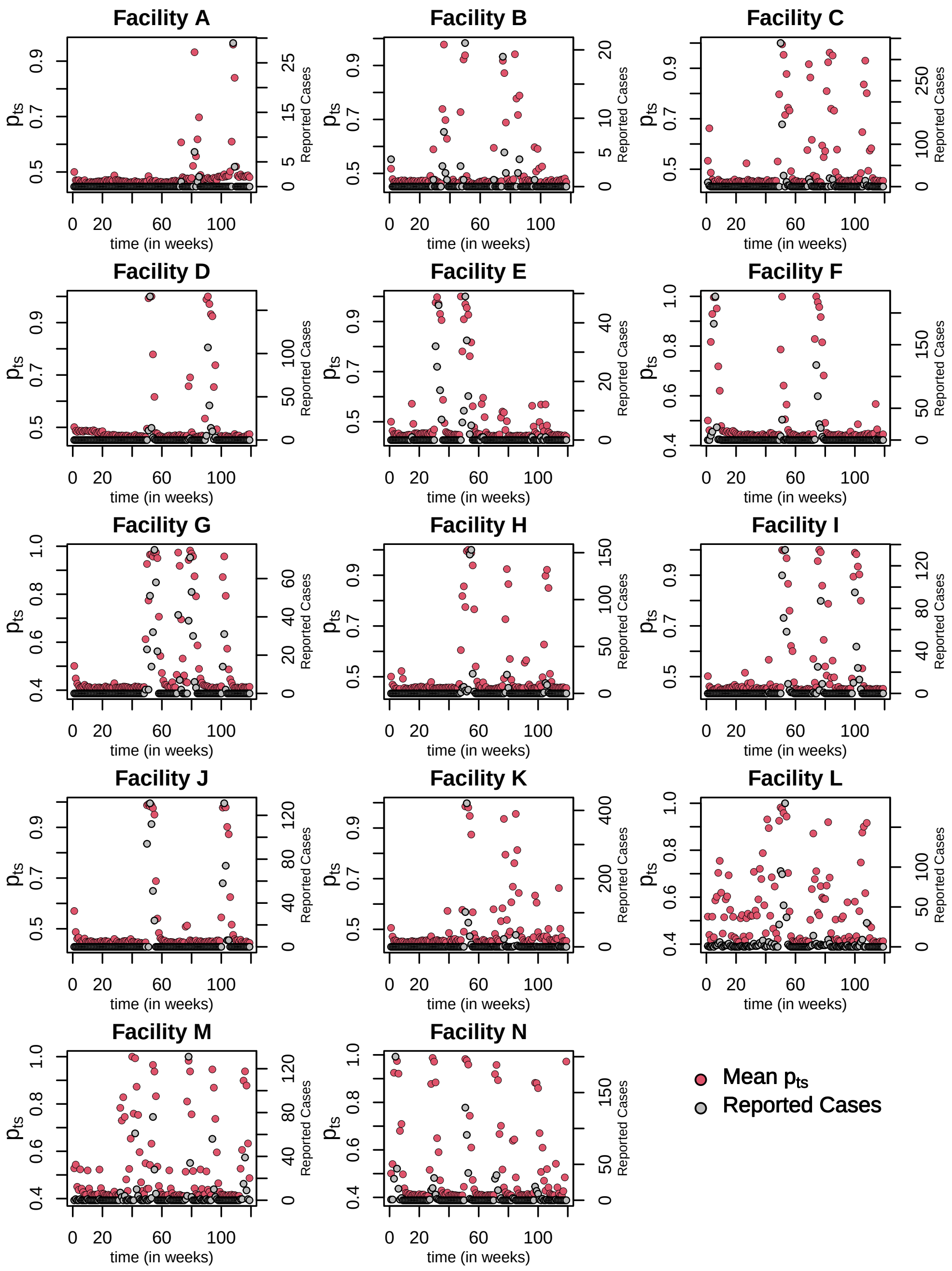

Figure J.12: Estimated $p_{ts}$ probabilities, the probability of at least one positive case versus the number of reported cases, $Z_{ts}$, by location under the SARS-CoV2-(N1) model.

Table J.12: Estimated SARS-CoV-2 (N1) ratio by location which can confirm one case with 80% probability.

| Facility | SARS-CoV-2 (N1) | | | Mean of $p_{ts} \geq 0 \cdot 8$ |
|---|---|---|---|---|
| | Minimum | Median | Mean | |
| A | 436·896 | 1171·20 | 1171·20 | 0·94510 |
| B | 47·895 | 1454·49 | 1843·07 | 0·89269 |
| C | 353·786 | 2045·49 | 3010·78 | 0·90346 |
| D | 576·888 | 5380·72 | 7202·27 | 0·95318 |
| E | 95·763 | 767·07 | 3011·25 | 0·93699 |
| F | 64·695 | 1830·59 | 2929·55 | 0·92828 |
| G | 69·736 | 515·55 | 766·13 | 0·92861 |
| H | 26·580 | 426·07 | 2110·06 | 0·91028 |
| I | 44·017 | 2055·15 | 5341·16 | 0·94265 |
| J | 29·530 | 915·04 | 1526·76 | 0·98030 |
| K | 96·467 | 793·73 | 1283·44 | 0·93313 |
| L | 124·707 | 1686·08 | 2539·96 | 0·91896 |
| M | 56·818 | 1018·83 | 3449·29 | 0·91485 |
| N | 48·763 | 924·80 | 1457·87 | 0·93473 |
